# Prospective Population-Scale Validation of an Electronic Health Record–Based Model for Pancreatic Cancer Risk

**DOI:** 10.64898/2026.04.11.26350318

**Authors:** Eric Lahtinen, Nicole Shigiltchoff, Kai Jia, Steven Kundrot, Matvey B. Palchuk, Jeff Warnick, Lawrence Chan, Irving D. Kaplan, Mandeep Sawhney, Martin Rinard, Limor Appelbaum

## Abstract

**Background and aims:** Pancreatic ductal adenocarcinoma (PDAC) surveillance is limited to individuals with familial or genetic risk although most future cases arise outside these groups. In a retrospective study, PRISM, an electronic health record (EHR)-based PDAC risk model, identified individuals in the general population at elevated near-term risk of PDAC. We aimed to prospectively evaluate whether PRISM can identify high-risk individuals beyond current surveillance groups across U.S. health systems.

**Methods:** We performed a prospective multicenter cohort study after deployment of PRISM in April 2023 across 44 U.S. health care organizations. Eligible adults aged ≥40 years without prior PDAC received a single baseline risk score and were assigned to prespecified risk tiers. Patients were followed for incident PDAC for 30 months. We estimated tier-specific 30-month cumulative incidence (positive predictive value, PPV), number needed to screen (NNS), standardized incidence ratios (SIRs), and time from deployment and first high-risk flag to diagnosis.

**Results:** Among 6,282,123 adults assigned a PRISM score, 5,058,067 had follow-up; 3,609 developed PDAC. The highest-risk tier had 30-fold higher PDAC incidence than the study population. At the SIR 5 threshold, 30-month cumulative incidence was 0.35% (NNS, 284.2); at SIR 16, 1.14% (NNS, 87.4); and at SIR 30, 2.19% (NNS, 45.7). Median time from deployment to PDAC diagnosis was 9.5 months, and median time from first high-risk flag to diagnosis at SIR 5 was 3.5 years. Shapley additive explanations (SHAP) analyses supported patient- and tier-level interpretability.

**Conclusions:** Prospective deployment of PRISM across multiple U.S. health care organizations identified individuals at elevated near-term risk for PDAC, with substantial risk enrichment and lead time before diagnosis. These findings support the real-world scalability and generalizability of EHR-based risk stratification for risk-adapted early detection. ClinicalTrials.gov identifier NCT05973331

## 1 Introduction

Pancreatic Duct Adenocarcinoma (PDAC) remains a major clinical challenge because its nonspecific early symptoms, low population incidence, and the absence of a validated, noninvasive screening test mean that most patients are diagnosed at an advanced, incurable stage. Only approximately 14% of PDAC cases are detected at an early stage [1], and overall 5-year survival remains approximately 13% [2], reflecting the limited effectiveness of treatment once disease is advanced. In contrast, outcomes are substantially better when PDAC is detected at a localized stage [3], and most long-term survivors are identified through surveillance programs for individuals deemed high risk on the basis of high genetic or familial risk [4]. However, inherited/familial predisposition accounts for only 5–10% of PDAC [5], meaning that current surveillance frameworks do not address over 90% of PDAC that arises outside established hereditary pathways and therefore have limited population-level impact on stage shift and survival. To improve survival at the population level, earlier detection strategies will need to expand beyond these groups.

EHR-based risk enrichment strategies that leverage routinely collected clinical data may provide a pragmatic pathway to expand screening eligibility beyond hereditary cohorts by systematically identifying individuals whose near-term risk exceeds actionable thresholds, thereby enriching for occult cancer and providing clinically meaningful lead time for downstream evaluation [6]. Multiple risk models have been proposed for specific populations, such as new-onset diabetes [7], and for the broader general population [8],[9] using longitudinal electronic health record (EHR) signals. However, translation of these approaches into routine care has remained limited.

Several barriers have constrained clinical adoption of PDAC risk models. Prospective evidence that model performance is maintained in real-world settings is limited [10, 11]. In addition, many studies do not report decision-relevant metrics, such as positive predictive value and number needed to screen, that are needed to select operational thresholds [6]. How risk scores should be operationalized into feasible downstream pathways also remains unclear. Threshold-linked decision metrics and tier-specific lead-time estimates are needed to define pragmatic follow-up strategies that fit real-world clinical capacity and target an actionable near-term window rather than lifetime risk. Concerns about false-positive predictions and downstream resource use also limit adoption, particularly when model outputs are not embedded within stepwise evaluation pathways. In practice, risk scores may serve as an initial triage step, followed by lower-burden confirmation (e.g., repeat clinical assessment or biomarker testing) before escalation to pancreas-protocol imaging or endoscopic evaluation. Clinical adoption also requires clinician and health-system trust in model predictions, which may be strengthened by patient-level interpretability that explains the factors contributing to an individual’s risk score. Computational and data-integration requirements can also be a barrier to scalable deployment across heterogeneous health systems [12], potentially limiting adoption to well-resourced settings and widening disparities [13]. Finally, few studies have demonstrated scalable deployment across diverse health systems and patient populations with explicit attention to generalizability and equity [12].

We previously developed PRISM, an electronic health record (EHR)–based machine learning model to predict incident pancreatic ductal adenocarcinoma (PDAC) 6–18 months before clinical diagnosis. In retrospective development and validation across 55 US health care organizations (HCOs), including academic medical centers, community hospitals, and outpatient clinics within the TriNetX federated network, PRISM showed good discrimination and risk stratification, using only routinely collected structured EHR data in a large, racially, ethnically, and geographically diverse U.S. population, with prespecified thresholds that identified small high-risk subgroups with substantially elevated PDAC incidence. Details of model development and retrospective validation have been reported previously [9]. Because we used the same PDAC ICD-based outcome definition and largely the same TriNetX federated network as in retrospective derivation, this prospective silent deployment provides a direct test of whether PRISM’s prespecified fixed-specificity operating points reproduce the same enrichment gradient in real-world follow-up. Unlike retrospective simulation, prospective evaluation is sensitive to real-world observability, censoring, and the timing of diagnosis capture relative to data refresh and longitudinal linkage.

Consistent with proposed frameworks for AI-based risk stratification and clinical decision support [14, 1, 15] our work has followed a staged trajectory from retrospective model development and validation, including evaluation of calibration, generalizability across sites and patient subgroups (including race/ethnicity and geography), and threshold-linked decision metrics (PPV and NNS) at prespecified operating points, to now large-scale “silent” prospective deployment with up to 30 months of follow-up.

This study was designed to prospectively validate PRISM in a large, diverse, federated multicenter network and achieve the following aims: (1) evaluate PRISM’s performance in prospective, real-world follow-up; (2) define clinically actionable operating points by quantifying threshold-specific decision metrics across risk tiers (including positive predictive value and number needed to screen); (3) estimate tier-specific lead time to define a near-term, high-yield window for screening; and (4) assess implementation feasibility at scale, including computationally practical deployment across heterogeneous health systems and patient-level interpretability to support integration into clinical workflows.

## 2 Methods

### 2.1 Study design, setting, and population

We conducted an observational, prospective cohort study of adults within 44 U.S. HCOs contributing to the TriNetX federated EHR network [16]. Although the original retrospective PRISM study included 55 U.S. HCOs, the prospective analysis was limited to 44 HCOs selected by TriNetX based on operational requirements for longitudinal follow-up, including stable patient identifiers and adequate data-refresh cadence. These organizations include academic medical centers, community hospitals, and outpatient care settings.

On April 21, 2023, we implemented a silent deployment of the PRISM PDAC risk prediction model across participating HCOs. For each eligible patient, PRISM generated an individual risk score for incident PDAC at deployment. Model outputs were not displayed to clinicians and did not trigger alerts or decision support, and therefore did not influence management decisions. The study protocol was registered at ClinicalTrials.gov (identifier: NCT05973331).

The study population comprised all patients scored by PRISM during the April 2023 deployment across the 44 participating HCOs. The first date on which a PRISM score was generated for a given patient was defined as the deployment date. Each patient was scored once. Baseline demographic and clinical characteristics were ascertained from EHR data available on or before the deployment date. Details of the underlying eligibility criteria and model input features used to generate PRISM scores have been described previously. The primary analysis cohort comprised patients with up to 30 months of observable follow-up after the deployment date (through November 5, 2025).

We included adults who met the prespecified PRISM deployment criteria and who received a PRISM risk score during the April 2023 deployment. Included individuals were at least 40 years of age with at least 45 days of EHR history prior to cohort entry, and had no history of pancreatic cancer in the EHR before the deployment date or, where tumor registry data were available, in the tumor registry in the absence of a corresponding PDAC diagnosis code in the EHR (see Supplement). Patients with a history of non-PDAC malignancies were not excluded.

### 2.2 Outcomes and follow-up

The primary outcome was incident PDAC within 30 months of the deployment date. Incident PDAC was defined as the first post-deployment occurrence of a qualifying PDAC ICD diagnosis code within the prespecified 30-month horizon (See Supplement). This single-code definition was prespecified and consistent with our prior PRISM study, in which stricter PDAC definitions did not materially change discrimination [17].

Secondary outcomes included the timing of incident PDAC occurrence, quantified as (i) the interval (lead time) between the date of deployment and the first PDAC diagnosis date and (ii) the interval from the first high-risk flag to the PDAC diagnosis date, with the first high-risk flag defined in Section 2.5; all-cause mortality within 30 months, defined using death indicators and recorded dates of death.

Vital status was ascertained using month- and year-level death dates, which we used to approximate time from deployment to death in time-to-event analyses.

### 2.3 PRISM prospective deployment and statistical analysis

Full modeling details for PRISM, including feature construction, algorithm selection, and retrospective validation, have been published previously [17]. In the present study, we prospectively deployed PRISM in a silent, non-interventional fashion across 44 U.S. health care organizations in the TriNetX network. At each participating organization, eligible adults who met the prespecified eligibility criteria received a PRISM risk score at deployment (April 21, 2023), based on the same feature specification as in the derivation study. Risk scores were computed within the federated environment but were not written back to local EHR systems.

The April 2023 scored cohort was followed longitudinally through subsequent TriNetX data refreshes over the prespecified 30-month risk horizon. Follow-up time was calculated from deployment to the earliest of PDAC diagnosis, death, last recorded clinical encounter in the EHR, or the administrative data cutoff for the 30-month horizon. Patients were then classified into four mutually exclusive follow-up categories: incident PDAC within 30 months, death without PDAC, PDAC-free with at least 30 months of follow-up, or censored before 30 months. Death ascertainment is as defined in Section 2.4.

Model performance was evaluated longitudinally by month after deployment at prespecified risk thresholds (fixed-specificity operating points spanning 85% to 99.9%) carried forward from the retrospective derivation analysis. For each month of follow-up, we generated performance tables summarizing AUC, sensitivity, specificity, PPV, NNS, and SIR, and timing metrics among incident PDAC cases: (i) the interval from deployment to PDAC diagnosis (MEPD) and (ii) the interval from earliest threshold crossing (“first high-risk flag”) to PDAC diagnosis (MEPDHiRisk). Because encounters before deployment were included, the first high-risk flag may occur before time 0 (negative months relative to deployment). For each patient, the first high-risk flag was defined as the earliest date on which the patient’s available EHR data would have produced a PRISM score at or above a prespecified threshold. Because PRISM was deployed once in April 2023 and scores were generated once per patient at deployment, each patient could contribute at most one first high-risk flag. Detailed follow-up and censoring rules are provided in the Supplementary Methods. Discrimination was quantified using AUC for incident PDAC within 30 months including time-dependent AUC estimates across months 1–30 of follow-up. Prespecified subgroup analyses evaluated discrimination (AUC) stratified by age, sex, race/ethnicity, and geographic region, consistent with the registered analysis plan (ClinicalTrials.gov NCT05973331). Because PRISM was locked at deployment, calibration was assessed without recalibration using reliability plots at 6, 12, 24, and 30 months and summarized using the geometric mean of the ratio of predicted to observed risk (Geometric Mean of Over Estimation; GMOE).

Although all thresholds are presented, for interpretability, we additionally express each operating point as an standardized incidence ratio (SIR), reflecting risk enrichment relative to the overall deployment cohort at the same month. We highlighted two reference operating points (SIR approximately 5 and SIR *≥* 16) for interpretability, not as prespecified clinical decision thresholds. For each threshold and each month of follow-up, we estimated sensitivity, specificity, positive predictive value (PPV), number needed to screen (NNS, the reciprocal of PPV), standardized incidence ratios (SIRs) and timing metrics (MEPD and MEPDHiRisk), as described above. Monthly PPV was estimated as the cumulative proportion of flagged patients diagnosed with PDAC by each month of follow-up. Missingness indicators were retained unchanged from the locked PRISM feature set, allowing the model to use informative missingness without prospective imputation; in descriptive analyses, missing race/ethnicity was reported as an “Unknown” category. Detailed definitions of PPV and SIR estimation and loss-to-follow-up handling, as well as additional implementation details, are provided in the Supplementary Methods. Statistical analyses were conducted using Python (Python Software Foundation).

### 2.4 Model interpretability

Model interpretability analysis was conducted using SHAP (SHapley Additive exPlanations) analysis, a game theory-based method for determining the marginal contribution of each feature across all possible feature combinations on model outcome [18]. For each patient, each feature was assigned a (SHAP) value reflecting its contribution to the PDAC risk score. All incident PDAC cases observed within 30 months of deployment in the prospective validation cohort were included in interpretability analyses (n=3,609). Summary (beeswarm) plots were generated within each prespecified risk-threshold (SIR) category to identify the key driving features for each risk group. We also generated patient-level SHAP waterfall plots for an example patient from each risk category to illustrate additive feature contributions underlying individual predictions. Example patients were selected by random sampling from each risk group. Technical implementation details are provided in the Supplementary Methods.

### 2.5 Ethical oversight

The data used in this study were collected between April 2023 and November 2025 from the TriNetX U.S. network, which provided access to de-identified EHRs from approximately 6.1 million patients across 44 participating HCOs. Our institutions maintain an individual, no-cost collaboration agreement with TriNetX, LLC that governs access to these data.

This observational cohort study used secondary analysis of existing EHR data and did not involve any intervention or interaction with human subjects. Data were de-identified in accordance with the HIPAA Privacy Rule and attested to by expert determination under Section §164.514(b)(1), most recently refreshed in December 2020. The Western Institutional Review Board determined that this research qualified as non–human subjects research.

This study followed the Strengthening the Reporting of Observational Studies in Epidemiology (STROBE) reporting guidelines.

## 3 Results

### 3.1 Study population and Baseline characteristics

Across 44 U.S. health care organizations, 6,282,123 adults met prespecified eligibility criteria and received a PRISM score during the April 2023 deployment window (Figure 1). The primary analysis cohort comprised 5,058,067 patients with observable follow-up through 30 months after the date. The remaining eligible patients did not have complete 30-month observability because of death or incomplete EHR follow-up (Figure 1). Within the primary analysis cohort, 3,609 patients were diagnosed with PDAC within 30 months (cumulative incidence, 0.07%). This observed occurrence was broadly consistent with expected rates for an adult population aged over 40 years [19].

**Figure 1:**
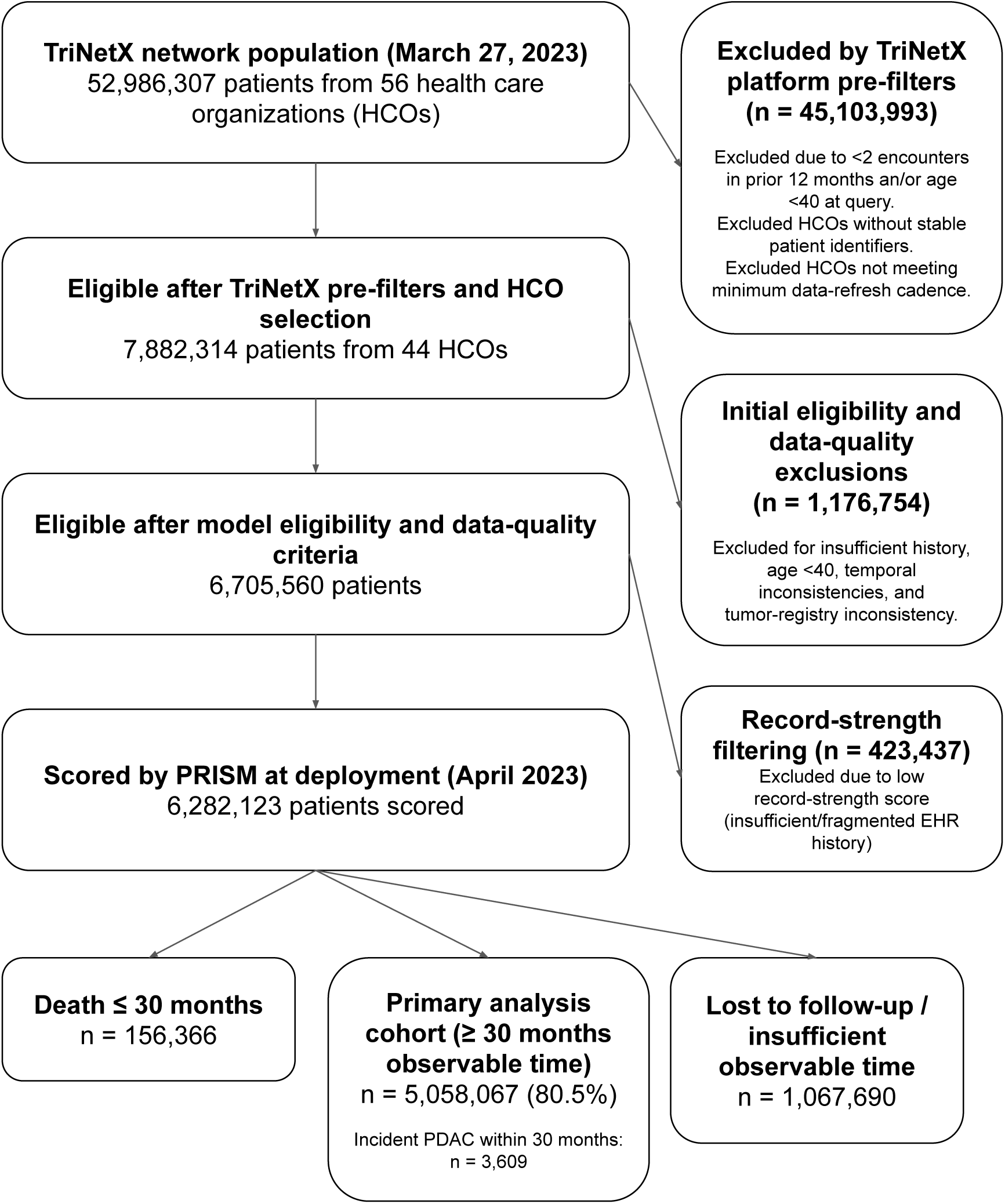
Flow (waterfall) diagram of the 30-month primary analysis cohort. After applying prespecified inclusion/exclusion criteria across 44 TriNetX health care organizations, 6,282,123 adults were eligible. After 30 months of follow-up, 5,058,067 (80.5%) remained with *≥*30 months of observable time and comprised the primary analysis cohort. Within this cohort, 3,609 developed pancreatic ductal adenocarcinoma (PDAC) within 30 months (cumulative risk 0.0709%). Stepwise counts and reasons for exclusion are shown in the diagram

Baseline characteristics of the primary analysis cohort are shown in Table 1. Of 6,282,123 patients, 3,609 developed PDAC within 30 months and 5,054,458 remained PDAC-free (Table 1). Cases were older (70.2±10.2 vs. 63.412.3 years) and more often male (48.6% vs. 41.9%). Among cases, 81.5% were aged 60 years or older, compared with 58.0% of non-cases. Race/ethnicity distributions were similar across groups (White: 74.0% vs. 72.2%; Black: 16.1% vs. 15.3%). Geographic distribution across health care organization regions was similar, with modest differences (e.g., Midwest 22.0% vs. 16.4%; Northeast 30.3% vs. 34.8%). Cases had greater EHR utilization (mean records, 349.2 ± 734.9 vs. 218.3 ± 501.2.).

**Table 1:** Baseline characteristics of the 30-month primary analysis cohort. (n=5,058,067), overall and by PDAC *≤*30m vs No PDAC *≤*30m with standardized mean differences (SMD).

| Attribute | | PDAC ≤ 30m (n=3,609) | | NO PDAC ≤ 30m (n=5,054,458) | |
| --- | --- | --- | --- | --- | --- |
|  |  | N | (%) | N | (%) |
| Sex | Female | 1,853 | (51.34) | 2,933,818 | (58.04) |
|  | Male | 1,756 | (48.66) | 2,120,166 | (41.95) |
|  | Unknown | 0 | (0.00) | 474 | (0.01) |
| Age at cutoff | Mean (SD) | 70.19 | (10.16) | 63.38 | (12.34) |
|  | 40 - 50 | 151 | (4.18) | 895,291 | (17.71) |
|  | 50 - 60 | 429 | (11.89) | 1,125,537 | (22.27) |
|  | 60 - 70 | 1,082 | (29.98) | 1,381,054 | (27.32) |
|  | 70 - 80 | 1,276 | (35.36) | 1,084,599 | (21.46) |
|  | > 80 | 583 | (16.15) | 463,263 | (9.17) |
|  | Unknown | 88 | (2.44) | 104,714 | (2.07) |
| Race | AIAN | 12 | (0.33) | 20,156 | (0.40) |
|  | Asian | 72 | (2.00) | 111,540 | (2.21) |
|  | Black | 581 | (16.10) | 774,762 | (15.33) |
|  | NHPI | 1 | (0.03) | 4,874 | (0.10) |
|  | White | 2,672 | (74.04) | 3,651,698 | (72.25) |
|  | Other Race | 0 | (0.00) | 0 | (0.00) |
|  | Unknown | 271 | (7.51) | 491,428 | (9.72) |
| HCO location | Midwest | 794 | (22.00) | 829,402 | (16.41) |
|  | Northeast | 1,091 | (30.23) | 1,759,267 | (34.81) |
|  | South | 1,417 | (39.26) | 1,865,541 | (36.91) |
|  | West | 272 | (7.54) | 534,620 | (10.58) |
|  | Unknown | 35 | (0.97) | 65,628 | (1.30) |
| No. medical records | Mean (SD) | 349.19 | (734.90) | 218.27 | (501.20) |

### 3.2 Discrimination and calibration

Discrimination was stable over follow-up, with time-dependent AUC estimates of AUC 0.740 (0.725,0.755) at 6 months, AUC 0.738 (0.727,0.748) at 12 months, AUC 0.739 (0.731,0.748) at 24 months, and AUC 0.742 (0.734,0.750) at 30 months. Calibration assessed without recalibration showed good agreement between predicted and observed risk across risk deciles on reliability plots at 6, 12, 24, and 30 months, with a 30-month GMOE of 1.078 (values greater than 1 indicate overestimation and values less than 1 indicate underestimation) (Figure 3).

### 3.3 Threshold-based operating characteristics and prospective yield

At prespecified operating points carried forward from derivation, higher thresholds flagged fewer patients and yielded greater PDAC enrichment (Table 2). At the operating point approximating SIR 5, PRISM flagged 254,643 patients (5.01% of the cohort) with a 30-month PPV of 0.35% (NNS, 284) and a mean time from first high-risk flag to diagnosis of 1,914 days. At the SIR 16.1 operating point, PRISM flagged 25,669 patients (0.505%) with a 30-month PPV of 1.15% (NNS, 87) and a mean lead time of 1,351 days, reflecting a more selective tier with higher yield. At the highest prespecified operating point (SIR 30.9; specificity 99.9%), PRISM identified an ultra–highrisk subgroup of 5,190 patients (0.102%) with a 30-month PPV of 2.21% (NNS, 45.3; and a mean lead time of 1,371 days (Table 2).

**Table 2:** Prespecified operating points and decision metrics at 30 months. PRISM was deployed in April 2023 in a silent, non-interventional manner across 44 TriNetX healthcare organizations. Among 5,058,067 eligible adults with 30-month observability including 3,609 incident PDAC cases, discrimination at 30 months was AUC 0.742 (95% CI, 0.734–0.750). Performance is shown at prespecified PRISM operating points carried forward from retrospective derivation corresponding to fixed-specificity thresholds (85%–99.9%). “Score cutoff” denotes the raw PRISM score threshold at each operating point, sensitivity reflects incident PDAC within 30 months; PPV reflects the observed 30-month cumulative incidence among flagged patients; NNS is the reciprocal of PPV; and SIR compares observed incidence in the flagged group with that in the overall study population. Timing metrics summarize time from first high-risk threshold crossing (“first high-risk flag”) to diagnosis (MEPD HiRisk) and time from deployment to diagnosis (MEPD Deploy). Values in parentheses are 95% CIs. Prespecified cutoffs do not represent observed 30-month risk. (See Table S2 for stratified results.)

| Score cutoff | Sens. | Spec. | PPV | NNS | SIR | MEPD<br>HiRisk(d) | MEPD<br>Deploy(d) |
| --- | --- | --- | --- | --- | --- | --- | --- |
| 7.8045 | 45.44% | 85.00% | 0.22% | 463.3 | 3.0 | 2173 (2043 to 2282) | 307 (291 to 325) |
| 7.2963 | 38.49% | 89.00% | 0.25% | 401.3 | 3.5 | 2074 (1974 to 2185) | 299 (281 to 319) |
| 6.6809 | 24.88% | 95.00% | 0.35% | 282.4 | 5.0 | 1914 (1818 to 2063) | 292 (270 to 311) |
| 6.4077 | 18.62% | 97.00% | 0.44% | 226.7 | 6.2 | 1766 (1658 to 1859) | 286 (258 to 307) |
| 5.4410 | 8.15% | 99.50% | 1.15% | 87.0 | 16.1 | 1351 (1209 to 1454) | 284 (236 to 310) |
| 4.6181 | 3.21% | 99.90% | 2.21% | 45.3 | 30.9 | 1371 (1218 to 1562) | 320 (231 to 385) |

At the same prespecified fixed-specificity operating points carried forward from the retrospective derivation, PRISM again produced a monotonic risk gradient in prospective follow-up (higher threshold *→* fewer patients flagged *→* higher PPV/SIR and lower NNS). At the operating point approximating SIR 5, the prospective 30-month PPV was 0.35% with SIR 5.0, compared with 0.58% and 5.10 retrospectively. At the prospective operating point approximating SIR 16.1 (specificity 99.5%), the 30-month PPV was 1.15% with SIR 16.1, compared with a retrospective PPV of 1.94% and SIR 19.8 at the closest retrospective tier (specificity 99.2%) (Table 2).

During silent prospective deployment, cumulative PDAC incidence among patients flagged at baseline increased over follow-up and was consistently higher at more selective operating points (Figure 3). The operating point approximating SIR 5 yielded modest enrichment with longer lead time, whereas higher thresholds identified smaller subgroups with progressively greater PDAC enrichment over time (Figure 2; Table 2).

**Figure 2:**
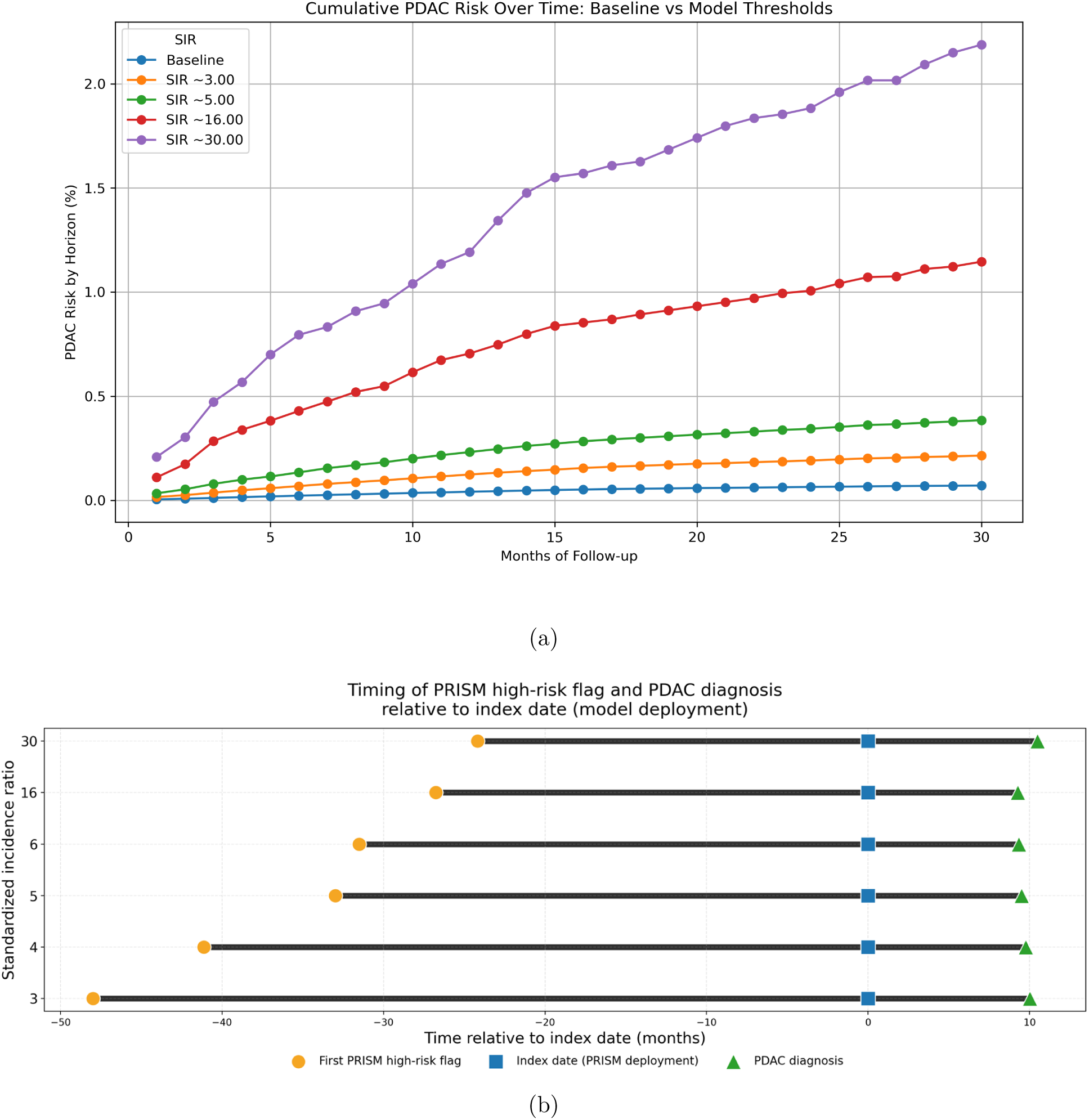
Prospective PRISM deployment: enrichment over time and lead time. **(a)** Cumulative PDAC incidence among flagged patients over follow-up. Monthly cumulative risk (PPV) over 30 months is shown for patients flagged at selected prespecified SIR tiers versus the baseline cohort incidence. Higher SIR tiers flag fewer patients with greater enrichment. Confidence intervals are omitted for legibility (see Table 2). (see Table 2). **(b)** Lead time relative to deployment and diagnosis at prespecified SIR tiers. For each SIR threshold defining high-risk status, points indicate (i) the first high-risk flag (circles), (ii) deployment date (squares; month 0), and (iii) the PDAC diagnosis date (triangles), all referenced to deployment. Horizontal segments connect first high-risk flag to diagnosis (MEPD HiRisk), summarizing time from positive prediction to diagnosis. Higher SIR tiers correspond to shorter intervals from high-risk flag to diagnosis.

**Figure 3:**
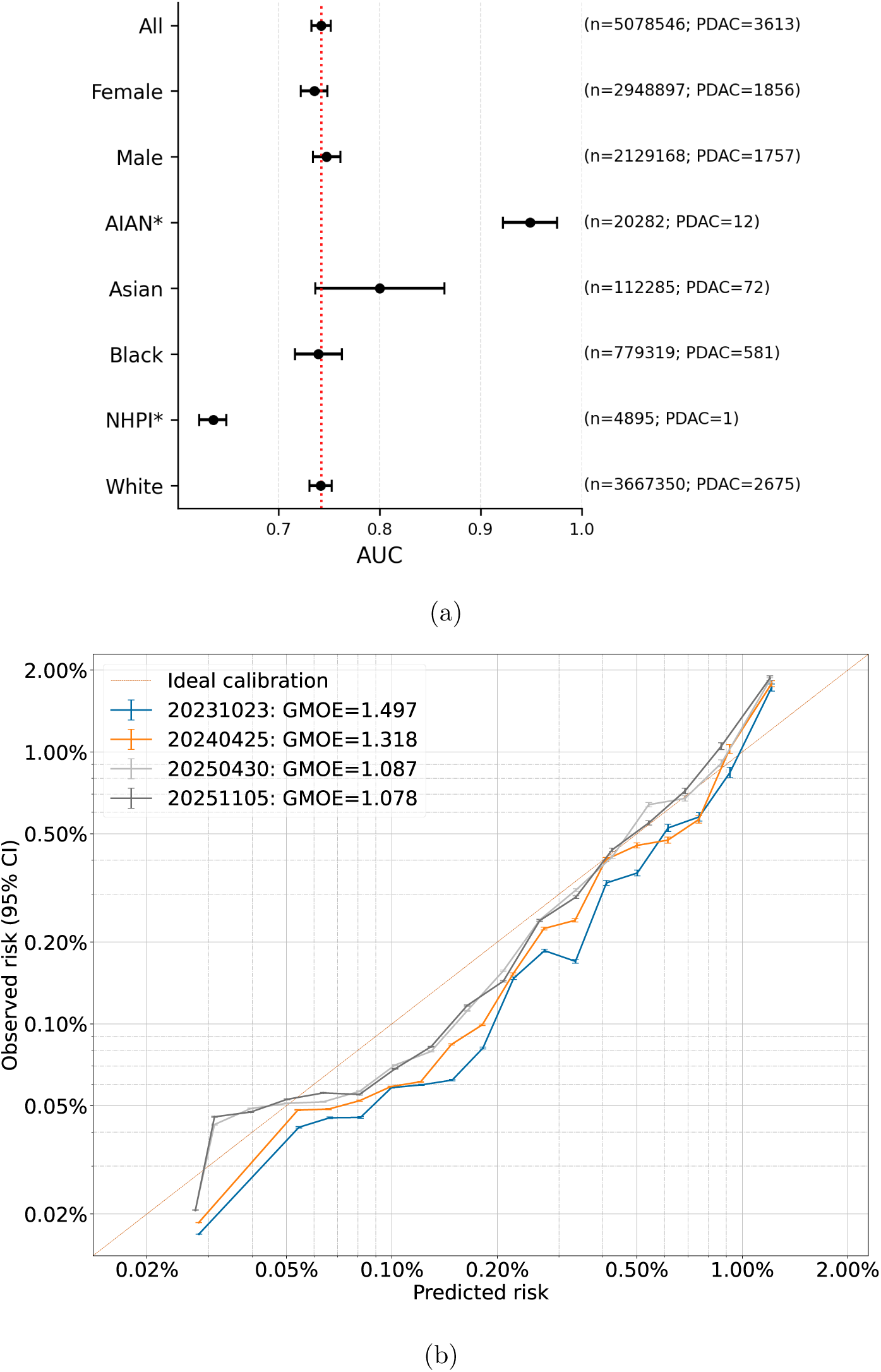
Prospective performance of PRISM in the overall cohort and key demographic subgroups. **(A)** Forest plot of discrimination (area under the receiver operating characteristic curve [AUC]) for the overall cohort and key demographic subgroups (sex and race/ethnicity). Points indicate AUC estimates and horizontal bars indicate 95% confidence intervals; subgroup sample sizes and incident PDAC case counts are shown. Estimates for subgroups with very small numbers of PDAC cases (e.g., AIAN and NHPI) should be interpreted cautiously due to greater uncertainty. **(B)** Calibration of PRISM at 6, 12, 24, and 30 months without recalibration (reliability plots). Predicted risk is plotted against observed event rates across prespecified risk bins; the 45° line indicates perfect calibration. Geometric Mean Over Estimate (GMOE) is shown for each group.

### 3.4 Subgroup performance

Discrimination was similar across prespecified demographic subgroups (Figure 3). Overall prospective performance was AUC 0.742 (95% CI, 0.733–0.752), with comparable discrimination in females (AUC, 0.735; 95% CI, 0.722–0.748) and males (AUC, 0.747; 95% CI, 0.734–0.761). Across the largest race and ethnicity strata, AUCs were similar with overlapping confidence intervals (White: 0.742 [95% CI, 0.731–0.753]; Black: 0.739 [95% CI, 0.716–0.762]; Asian: 0.800 [95% CI, 0.736–0.864]). Estimates in smaller strata (e.g., NHPI and AIAN) were imprecise because of limited numbers of incident PDAC cases and should be interpreted cautiously.

Calibration was qualitatively similar across subgroups on reliability plots (Supplementary Appendix, Figure S2).

### 3.5 Model interpretability

Across higher-risk SIR strata, the strongest contributors by absolute SHAP magnitude included age, kidney function markers (eGFR/creatinine), and healthcare utilization (encounter frequency), with additional contributions from glycemic/diabetes-related features and selected pancreatic, biliary, and symptom codes (Figure 4 A-C). At SIR 5, in addition to the dominant effects of age, kidney function, and utilization, glycemic markers (HbA1c/eAG) and GI/abdominal symptom and neoplasm-related codes (e.g., abdominal pain; digestive-system neoplasm diagnoses) contributed notably. At SIR approximately 16, additional contributors included glycemic markers (glucose/HbA1c/ eAG) and select hematologic/liver tests (e.g., RDW, direct bilirubin) and symptom codes (e.g., abdominal pain) (Figure 4 B-C).

**Figure 4:**
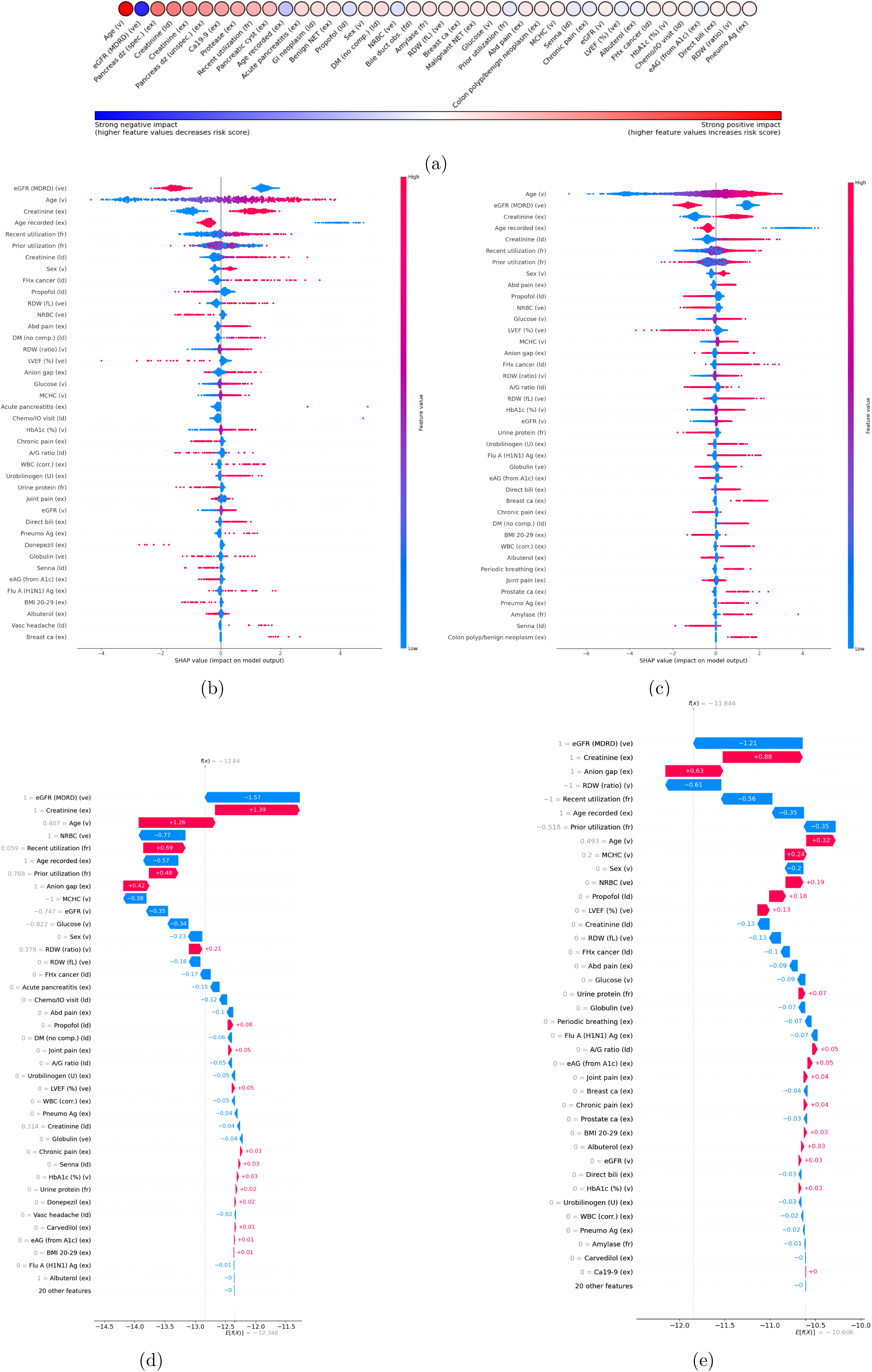
SHAP feature contributions for PRISM: (a) Global feature importance for the 40 features with the largest mean absolute SHAP values, ranked by mean SHAP value. Red indicates features for which higher values increase the model output and therefore predicted PDAC risk, whereas blue indicates features for which higher values decrease the model output. Color intensity reflects effect magnitude. (b-c) Beeswarm plots showing the distribution and direction of feature contributions among patients flagged at SIR 5 and SIR 16. Each point represents a patient-feature pair; color indicates the feature value (red = higher values, blue = lower values), and horizontal position reflects the SHAP value (contribution to the predicted PDAC risk). Features are ordered by mean SHAP value within each risk stratum, (d-e) Waterfall plots for a randomly selected patients from the SIR 5 and SIR 16 risk group. Red arrows increase the SHAP value and therefore risk score relative to the cohort mean, blue arrows decrease the SHAP value and therefore risk score relative to the cohort mean. Longer arrows = stronger effect. The feature value is in gray to adjacent to the feature label.

Waterfall plots for representative patients in the SIR 5 and SIR 16 strata illustrated patient-level heterogeneity in risk attribution, with individual predictions reflecting different combinations of familiar clinical features and less intuitive EHR-derived signals (Figure 4 D-E).

## 4 Discussion

We report a large-scale, prospective silent deployment of PRISM, an EHR-based PDAC risk model, across 44 U.S. health care organizations in the TriNetX network in more than 6 million adults, with 30 months of follow-up. Using the same prespecified fixed-specificity operating points carried forward from our retrospective work, the prospective deployment reproduced the expected risk gradient with higher thresholds flagging fewer patients and yielding higher PPV/SIR and lower NNS. Absolute enrichment at the highest tiers was attenuated relative to retrospective simulated estimates, consistent with real-world prospective follow-up. The principal findings are as follows: (1) PRISM enriched the general population, identifying subgroups with up to 30-fold higher near-term PDAC risk and providing tier-specific lead time, with a median 3.5 years from first high-risk flag to diagnosis (SIR 5) and 9.5 months from deployment to diagnosis, defining an actionable window for risk-adapted downstream evaluation. These timing results suggest that PRISM captures earlier EHR patterns associated with near-term PDAC risk rather than only immediate pre-diagnostic signals; (2) PRISM demonstrated stable prospective performance over the follow-up period, with consistent discrimination (AUC 0.74 across months 1–30) and calibration across key demographic and health-system subgroups, and feasibility of population-scale deployment and generation of individual risk scores across a federated network; (3) Thirty-month prospective follow-up yielded real-world decision metrics at the same prespecified PRISM cutoffs used in the retrospective simulated deployment, converting individual risk scores into SIR-based tiers with observed PPV and NNS (PPV up to 2.19%; NNS 45.7); (4) interpretability analyses (SHAP) highlighted established clinical risk correlates and potentially novel signals associated with predicted risk, supporting face validity, generating testable hypotheses for future study, and providing patient-level explanations to accompany each risk score.

Despite progress in PDAC risk modeling, translation into routine care has remained limited. Pancreatic ductal adenocarcinoma remains a major clinical challenge because most patients are diagnosed at an advanced, incurable stage, underscoring the need to identify more individuals earlier in the disease course. Current PDAC surveillance programs primarily focus on inherited predisposition or strong family history. Inherited/familial risk accounts for only 5-10% of PDAC, meaning that existing surveillance frameworks do not address more than 90% of cases that arise outside established hereditary pathways. By enabling risk enrichment using routinely collected EHR data, PRISM has the potential to extend near-term risk stratification to this much larger population and thereby increase the potential population-level impact of earlier detection.

Although multiple risk models have been developed for specific populations (e.g., new-onset diabetes) [7] and for the broader general population [8, 9], several barriers have constrained clinical adoption. A key barrier is the need for prospective evidence that model performance is maintained in real-world settings, alongside demonstration that risk stratification can be applied at scale across diverse health systems and demographic subgroups without exacerbating disparities. Additional challenges include transportability across heterogeneous EHR coding and practice patterns, reliance on inputs that may be inconsistently measured in routine care, and uncertainty about how to translate risk scores into downstream actions (including the burden of false-positive workups). In addition, implementation requires operationally meaningful outputs, such as threshold-based decision metrics and lead time, so clinicians and health systems can select pragmatic cutoffs and define follow-up actions for newly identified high-risk patients. Bridging the gap between retrospective model development and prospective evaluation in real-world workflows is therefore a central prerequisite for clinical translation.

As recently noted, computational considerations, including the complexity of EHR extraction and preprocessing, runtime, and pipeline scalability, often determine whether a risk model can be realistically integrated into clinical workflows, independent of statistical performance [12]. Our study directly addresses these implementation challenges. In partnership with TriNetX, we operationalized a standardized, scalable scoring pipeline across 44 health care organizations and generated individual risk scores for over 6 million adults at deployment with prospective follow-up over 30 months, demonstrating the computational feasibility of prospective evaluation at population scale. In our deployment, model execution and scoring were not a practical bottleneck. Instead, the dominant operational dependency was the cadence of upstream EHR data refresh, harmonization, and quality control across participating health systems, which were performed within the TriNetX federated platform, allowing large-scale scoring to be executed efficiently across 44 organizations. In this context, federated EHR networks offer a practical pathway for population-scale prospective evaluation and potential implementation by enabling standardized, harmonized analyses across multiple health systems while keeping data local. The present silent deployment leverages this infrastructure to assess PRISM prospectively on a common evaluation platform across 44 organizations, providing evidence of maintained performance and calibration over time, including across demographic and geographic subgroups. Beyond discrimination, we report operationally interpretable outputs at prespecified thresholds: PPV, NNS, and standardized incidence ratios, along with prospective timing metrics that quantify the lead-time window available between high-risk identification and diagnosis. Together, these results move beyond retrospective accuracy by demonstrating real-world feasibility and by providing the threshold-linked information needed to design pragmatic workflows (e.g., biomarker triage, imaging, or referral pathways) and future interventional studies. Finally, PRISM outputs both a patient-level risk score and transparent patient-level explanations (e.g., waterfall-style SHAP decompositions), which may support clinician interpretation and communication at the point of care and help set the stage for more personalized approaches to PDAC surveillance in which screening eligibility, timing, and intensity are adapted to near-term risk.

To support clinical interpretation, we mapped prespecified fixed-specificity operating points to standardized incidence ratios (SIRs), allowing conceptual comparison to how “high-risk” is defined in current PDAC surveillance frameworks [20]. However, guideline eligibility is fundamentally anchored to lifetime predisposition, whereas PRISM is designed to identify near-term risk over a fixed horizon. In this prospective silent deployment, a SIR 5 operating point—chosen to parallel the commonly cited “5-fold risk” concept—identified approximately one quarter of PDAC cases occurring within 30 months (sensitivity 24.8%), extending risk stratification beyond the relatively small proportion of PDAC attributable to familial/hereditary predisposition. Importantly, higher-risk tiers identified by PRISM achieved near-term risk levels comparable to those observed in established surveillance cohorts. For example, in the CAPS multicenter screening study, PDAC incidence was approximately 5.15 cases per 1,000 person-years—equivalent to roughly one PDAC diagnosis per 194 individuals screened per year, a yield similar to that observed in the PRISM SIR 16 tier (NNS approximately 87 over 30 months, or approximately 1 per 218 individuals per year). For context, this corresponds to an absolute risk of 1.14% over 30 months (1 in 88), which is 30-fold higher than the age-adjusted U.S. population incidence (13.8 per 100,000 person-years). In parallel, published economic analyses of PDAC surveillance suggest that cost-effectiveness is most favorable at higher-risk levels, supporting evaluation of more stringent tiers (e.g., SIR 16; sensitivity about 8%) as a “very-high-risk” option for more intensive downstream evaluation, while lower tiers (e.g., SIR 5) may be better suited to stepwise, resource-conserving pathways that can be prospectively tested in pragmatic implementation studies.

A practical implication of these findings is that PRISM could support a risk-adapted approach to PDAC evaluation rather than a single binary “screen/not screen” decision. Lower-tier thresholds (e.g., SIR 5) may be best suited to stepwise pathways that prioritize confirmatory assessment before advanced imaging, including laboratory/biomarker triage or referral to a high-risk clinic, whereas higher-tier thresholds (e.g., SIR 16 or greater) identify smaller groups with higher nearterm risk that may justify more intensive downstream evaluation, such as pancreas-protocol imaging and/or EUS-based pathways. In clinical practice, patients identified as high risk could potentially be considered for surveillance approaches similar to those used for individuals with hereditary or familial susceptibility to PDAC, in whom EUS and MRI are often alternated annually, while very-high-risk groups (e.g., SIR 30) may warrant consideration of shorter intervals in selected cases. Importantly, the observed lead-time window suggests that risk stratification could help time evaluation during a period of maximal near-term risk, but the optimal actions, thresholds, and frequency of follow-up will require prospective interventional studies and should ultimately be determined by treating physicians, taking into account patient comorbidities and preferences. More broadly, the practice of identifying individuals at elevated cancer risk and enrolling them in structured surveillance pathways is becoming increasingly accepted, particularly as early-detection strategies, including multi-cancer early detection approaches, continue to evolve.

This study has several limitations. First, PRISM was deployed in a silent, observational manner. Accordingly, we could not determine whether clinician-facing alerts would change diagnostic evaluation, timeliness of workup, stage at diagnosis, or patient outcomes. Second, risk scores were generated once at deployment, without longitudinal re-scoring, so we could not assess whether repeated scoring would identify additional patients earlier or improve operating characteristics. Third, there is no established benchmark for near-term PDAC risk thresholds in the general population: current surveillance frameworks are largely defined using lifetime risk in hereditary and familial-risk cohorts, limiting direct comparability and complicating selection of an “optimal” cutoff for broader implementation. Fourth, large-scale deployment, scoring, and longitudinal follow-up in this study were enabled by the TriNetX federated platform. Implementation outside such an environment may differ across institutions and may require site-specific calibration. Finally, although PDAC diagnosis was supplemented with tumor registry linkage where available, registry outcomes were available only for a minority of participating institutions. Thus, we could not directly assess whether PRISM preferentially identified patients before early-stage PDAC diagnosis. However, PRISM identified patients months to years before PDAC diagnosis overall, defining a lead-time window that is consistent with earlier identification and supports future evaluation of stage shift in studies with more complete stage ascertainment. In addition, we did not evaluate downstream diagnostic pathways, including utilization and costs associated with follow-up testing, which will be essential to assess in pragmatic prospective studies that integrate PRISM into real clinical workflows.

In conclusion, this 30-month silent prospective deployment shows that PRISM can identify subgroups at elevated near-term risk for PDAC and can be executed at population scale across a large, diverse U.S. population. PRISM also provides patient-level explanations to accompany each risk score, which may support clinical interpretation and future implementation. These findings provide a foundation for pragmatic clinician-facing studies testing whether risk-adapted pathways can expand surveillance beyond traditional hereditary and familial-risk populations.

## Data Availability

Deidentified individual participant data will not be shared directly by the authors. The data underlying this study are part of the TriNetX federated network, and access is governed by TriNetX and
participating health care organizations. Researchers interested in accessing these data may do so
only through separate agreements with TriNetX and the relevant institutions, subject to applicable
approvals and terms. The analytic methods are described in detail in the manuscript to support
reproducibility. Investigators interested in evaluating PRISM through the TriNetX platform may
contact the corresponding authors to discuss potential evaluation through the network.

## Author contributions

Conceptualization, LA, MR, and SK; Methodology, LA, MR, EL, SK, and KJ; Software, KJ, EL; Data curation, MR, LA, and EL; Validation, EL, NS, MR, and LA; Formal analysis, EL, NS, LC, MR, LA, MP, IDK, and MS; Investigation, JW, LC, EL, and NS; Resources, JW, LC, SK, and MP; Project administration, LA, MR, and JW; Visualization, EL, NS, and LA; Supervision, MR, LA, SK, and MP; Funding acquisition, LA; Writing – original draft, LA, EL, and NS; Writing – review and editing, all authors. All authors approved the final manuscript and agree to be accountable for all aspects of the work.

## Grant support

This work was supported in part by the generous philanthropy of Jane O. Siegel, DMD, and by TriNetX. TriNetX provided in-kind support, including secured laptop computers, access to the TriNetX EHR database, and technical, legal, and administrative assistance from its team of clinical informaticists, engineers, and technical staff. S.K., M.P., and J.W. are employees of TriNetX and were involved in study design and data acquisition. L.C. was an employee of TriNetX at the time this work was conducted. No formal grant funding supported this work.

## Acknowledgments

The authors are deeply grateful to Gadi Lachman, who served as Chief Executive Officer of TriNetX during most of the prospective validation period, for his strong and sustained support of this work. The authors are also grateful to Jason North for his instrumental role in establishing and launching the prospective validation study at TriNetX.

The authors gratefully acknowledge the generous support of Jane O. Siegel, DMD. The authors thank Gadi Lachman, Justin North, and the TriNetX team for their support of this work, including assistance in initiating the prospective validation effort and operational support throughout the study.

## Abbreviations

AIAN: American Indian or Alaska Native
eAG: estimated average glucose
eGFR: estimated glomerular filtration rate
GMOE: geometric mean of overestimation
HCO: health care organization
MEPD: mean elapsed time from deployment to diagnosis
MEPDHiRisk: mean elapsed time from first high-risk flag to diagnosis
NHPI: Native Hawaiian or Pacific Islander
NNS: number needed to screen
PDAC: pancreatic ductal adenocarcinoma
PPV: positive predictive value
PRISM: pancreatic cancer risk model
SHAP: SHapley Additive exPlanations
SIR: standardized incidence ratio
STROBE: Strengthening the Reporting of Observational Studies in Epidemiology.

## Disclosures

E.L., K.J., N.S., I.K., and L.A. report no conflicts of interest relevant to this work. M.S. reports stock ownership in Allurion Inc. and serving as principal investigator on a training grant from Boston Scientific, both outside the submitted work. S.K., M.P., and J.W. are full-time employees of TriNetX, LLC, and own stock in TriNetX. L.C. was an employee of TriNetX, LLC, during the conduct of this work. M.R. reports consulting, advisory, equity, research support, and patent relationships outside the submitted work, as detailed in the disclosure forms.

## Data transparency statement

Deidentified individual participant data will not be shared directly by the authors. The data underlying this study are part of the TriNetX federated network, and access is governed by TriNetX and participating health care organizations. Researchers interested in accessing these data may do so only through separate agreements with TriNetX and the relevant institutions, subject to applicable approvals and terms. The analytic methods are described in detail in the manuscript to support reproducibility. Investigators interested in evaluating PRISM through the TriNetX platform may contact the corresponding authors to discuss potential evaluation through the network.

## Declaration of Generative AI and AI-assisted technologies in the writing process

During the preparation of this work the authors used ChatGPT (OpenAI) in order to assist with editing and improving the clarity, grammar, style, and overall language of the manuscript. After using this tool/service, the authors reviewed and edited the content as needed and take full responsibility for the content of the publication.

## Notes

### Clinical Trial

NCT05973331

### Funding Statement

This work was supported in part by a philanthropic gift from Jane O. Siegel, DMD, and by TriNetX.
TriNetX provided in-kind support, including secured laptop computers, access to the TriNetX EHR
database, and technical, legal, and administrative assistance from its team of clinical informaticists,
engineers, and technical staff. S.K., M.P., and J.W. are employees of TriNetX and were involved
in study design and data acquisition. L.C. was an employee of TriNetX at the time this work was
conducted. No formal grant funding supported this work.

### Author Declarations

The data used in this study were collected between April 2023 and November 2025 from the TriNetX U.S. network, which provided access to deidentified EHRs from approximately 6.1 million patients across 44 participating HCOs. Our institutions maintain an individual, no-cost collaboration agreement with TriNetX, LLC that governs access to these data. This observational cohort study used secondary analysis of existing EHR data and did not involve any intervention or interaction with human subjects. Data were deidentified in accordance with the HIPAA Privacy Rule and attested to by expert determination under Section 164.514(b)(1), most recently refreshed in December 2020. The Western Institutional Review Board determined that this research qualified as non human subjects research.

