## Supplementary Materials for "Prospective Population-Scale Validation of an Electronic Health Record–Based Model for Pancreatic Cancer Risk"

#### Methods

#### PRISM Prospective deployment and performance assessment

**Follow-up** During the April 2023 implementation window, PRISM was executed automatically in the background to generate and store a PDAC risk score for each eligible patient. Patients were followed longitudinally through subsequent TriNetX data refreshes to ascertain outcomes over the prespecified 30-month risk horizon. TriNetX maintained persistent, de-identified patient identifiers, enabling longitudinal linkage of patients across repeated data extracts.

Following the initial April 2023 deployment, we obtained updated de-identified patient-level datasets from TriNetX approximately every 6 months. TriNetX performed longitudinal linkage across data refreshes using stable, de-identified patient identifiers (unique within each participating HCO), allowing follow-up of the same scored patients over the 30-month observation period. We did not re-define or rebuild the cohort at each timepoint. Instead, the April 2023 scored cohort was treated as a fixed prospective cohort, and subsequent downloads were used solely to update follow-up and outcome status for those same individuals.

For each patient in the April 2023 cohort, we recorded the deployment date (date of PRISM scoring), the corresponding PRISM risk score, and all subsequent PDAC diagnoses, deaths, and encounters. Death ascertainment was as defined in Section 2.4. Follow-up time was calculated from the deployment date to the earliest of PDAC diagnosis, death, last recorded clinical encounter in the EHR (used as the last-contact date), or the administrative data cut date for the 30-month horizon.

Using the most recent download (approximately 30 months after deployment), we classified each patient from the original 2023 cohort into one of four mutually exclusive categories: (1) developed PDAC within 30 months of the date; (2) died within 30 months without a recorded PDAC diagnosis; (3) alive and PDAC-free with at least 30 months of documented follow-up; or (4) censored before 30 months without PDAC (lost to follow-up). These classifications were derived from patient-level dates ( date, PDAC diagnosis date, death date, and last contact date).

**Event ordering** Patients were classified based on the first observed event within 30 months of the date. PDAC was counted if a PDAC diagnosis occurred before death (regardless of subsequent death), whereas death was counted if it occurred before any recorded PDAC diagnosis. Patients without PDAC who lacked 30 months of observable follow-up were censored at the last recorded encounter.

Model performance was evaluated longitudinally by month after the deployment date at prespecified risk thresholds (fixed-specificity operating points spanning 85% to 99.9%) carried forward from the retrospective derivation analysis. For each month of follow-up, we generated performance tables summarizing discrimination and classification metrics for the PRISM score, including AUC, sensitivity, specificity, PPV, NNS, and SIR, and timing metrics among incident PDAC cases: (i) the interval from deployment to PDAC diagnosis (MEPD) and (ii) the interval from earliest encounter date at which the PRISM score exceeded a given threshold ("first high-risk flag") to the PDAC diagnosis date (MEPDHiRisk); because encounters prior to the date were included for this timing analysis, the first high-risk flag may occur before time 0 (negative months relative to deployment). For each patient, we identified the first high-risk flag as the earliest date on which the patient's available EHR data would have produced a PRISM score at or above a prespecified threshold (based on the locked model and feature set). Because PRISM was deployed once in April 2023 and scores were generated once per patient at deployment, each patient could contribute at most one first high-risk flag. For some patients, the EHR data supporting threshold crossing were already present before deployment. In these cases, the first high-risk flag preceded deployment.

### 1 Statistical analysis

**PPV (cumulative incidence) estimation** For each prespecified threshold, we estimated monthly PPV as the cumulative proportion of flagged patients diagnosed with PDAC by each month of follow-up, calculated as the number of incident PDAC diagnoses observed through that month divided by the total number of patients flagged at baseline. Patients without PDAC who were lost to follow-up before 30 months contributed follow-up until their last recorded encounter. PPV estimates therefore reflect observed outcomes in the available EHR follow-up.

Patients lost to follow-up were not removed from the PPV denominator, therefore, PPV estimates are based on observed outcomes under incomplete EHR follow-up and may underestimate risk if additional PDAC diagnoses occurred after the last recorded encounter. Monthly PPV was estimated as the cumulative proportion of flagged patients diagnosed with PDAC by each month of follow-up. Because patients lost to follow-up were not removed from the PPV denominator, PPV estimates may underestimate risk if additional PDAC diagnoses occurred after the last recorded encounter. Detailed definitions of PPV and SIR estimation and missing-data handling are provided in the Supplementary Methods.

**SIR calculation** Standardized incidence ratios (SIRs) were calculated at each month of follow-up for each prespecified PRISM threshold as the cumulative PDAC incidence (PPV) among patients above the threshold divided by the cumulative PDAC incidence in the overall deployment cohort ("population risk") at the same month.

**Missing data** Missingness indicators (feature "existence bits," coded 0/1 for whether an EHR entry type was present) were included in the original PRISM feature set and retained unchanged in the locked prospective deployment, allowing the model to use informative missingness without prospective imputation. In descriptive analyses, missing race/ethnicity was reported as an "Unknown" category.

### 2 Interpretability

SHAP value calculations were done with the `GradientExplainer` algorithm from Python's SHAP library, with 50 background samples selected randomly. The SHAP version used was 0.48.0, and was run on Python 3.12.2 on AWS EC2 instance of type r6a.4xlarge. Beeswarm plots were generated using the `summary_plot` function, and waterfall plots were generated using the `waterfall_plot` function.

### 3 Outcome definition

In prior PRISM development analyses, we evaluated stricter PDAC definitions requiring either (i) $\geq$2 PDAC ICD-coded encounters or (ii) 1 PDAC ICD-coded encounter plus tumor registry confirmation. These stricter definitions reduced the number of PDAC cases but did not meaningfully change model discrimination. Therefore, we retained the single-code PDAC definition for the present prospective validation.

#### ICD diagnoses groupings

**PDAC Diagnosis Target Codes ICD-10:**

- C25.0
- C25.1
- C25.2
- C25.3
- C25.7
- C25.8
- C25.9

**Diagnosis Target Codes ICD-9:**

- 157
- 157.0
- 157.1
- 157.2
- 157.3
- 157.7
- 157.8
- 157.9

#### Figures

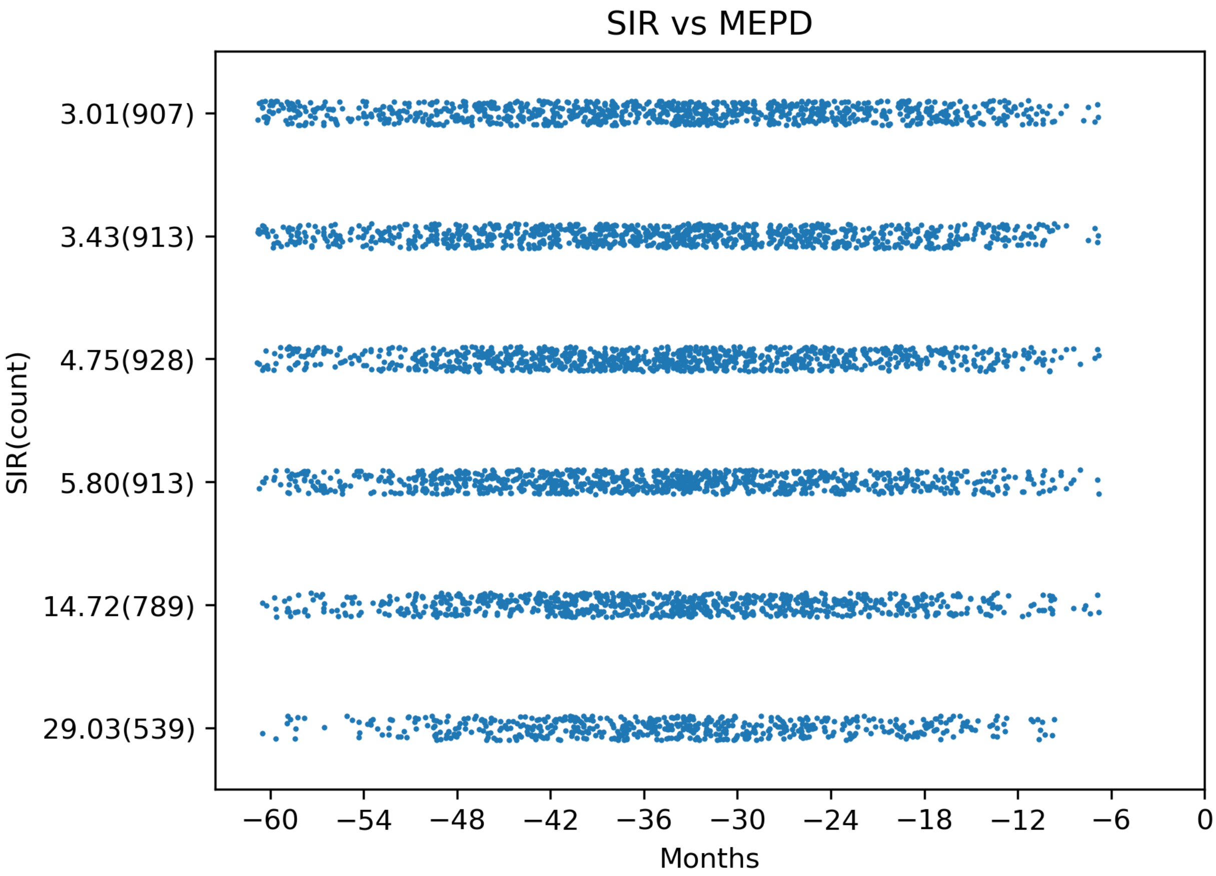


Figure S1: **Pre-deployment lead time from first high-risk flag** Beeswarm plot showing the pre-deployment lead time only: the distribution of months from the first high-risk flag to deployment/ (0 months) for each SIR threshold. Negative values indicate that the HR signature was present months to years before deployment. SIR labels include the number of PDAC cases contributing to each stratum.

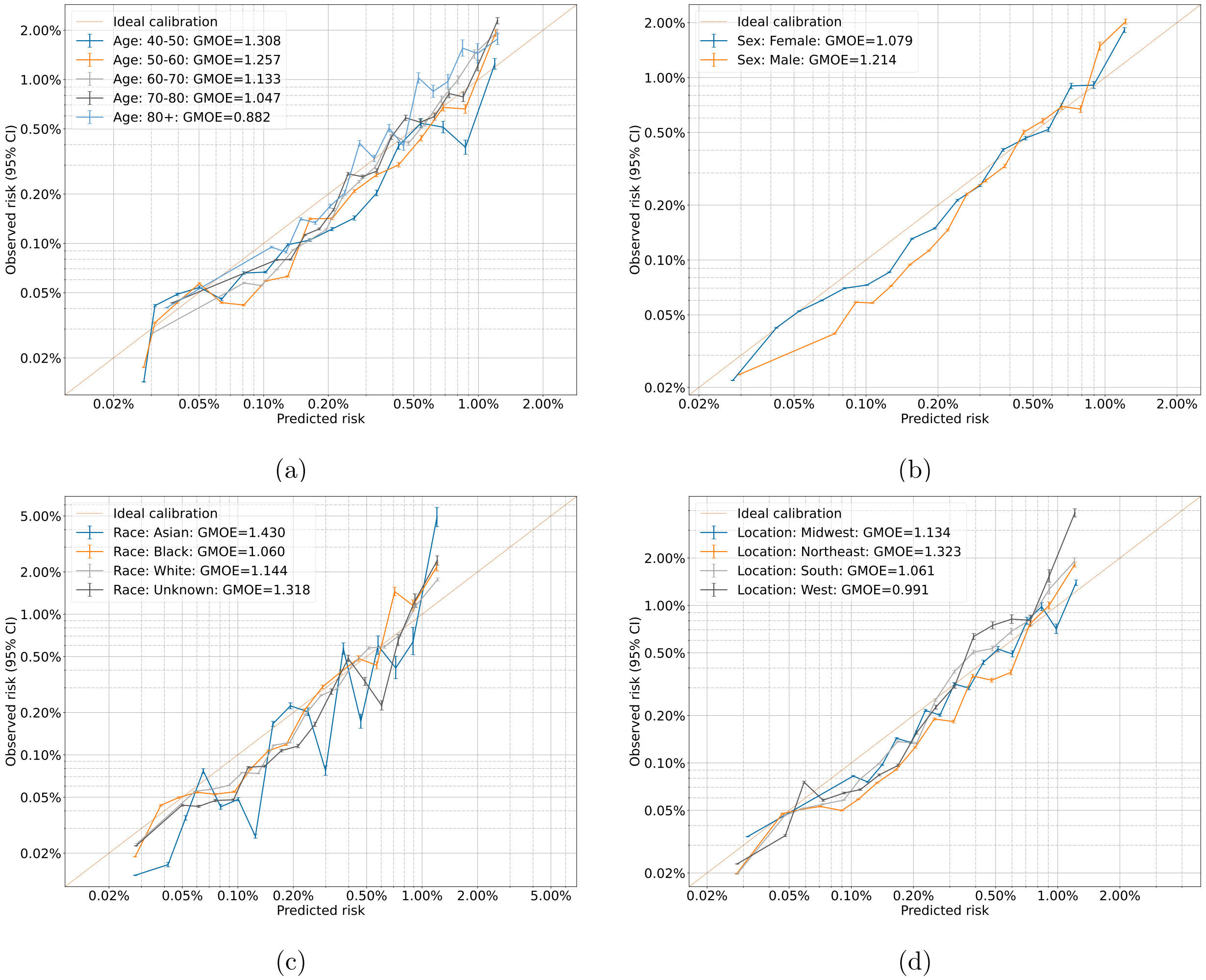


Figure S2: **Calibration curves by age, sex, location, and race**: Predicted risk is plotted against observed event rates across prespecified bins; the 45° line indicates perfect calibration. Geometric Mean Over Estimate (GMOE) is shown for each group. Smaller numbers in each bin size were related to worse calibration outcomes (see figure 2). AIAN, NHPI, unknown sex, and unknown location calibration curves were excluded due to very small data sample resulting in a very large GMOE.

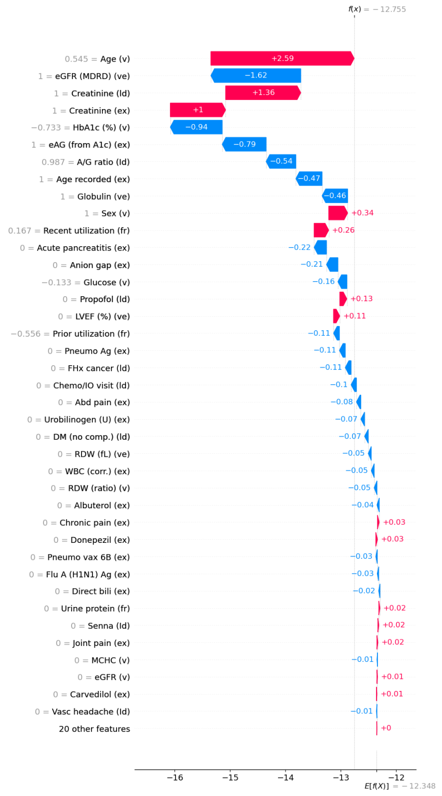
f(x) = −12.755
0.545 = Age (v) +2.59
1 = eGFR (MDRD) (ve) −1.62
1 = Creatinine (ld) +1.36
1 = Creatinine (ex) +1
−0.733 = HbA1c (%) (v) −0.94
1 = eAG (from A1c) (ex) −0.79
0.987 = A/G ratio (ld) −0.54
1 = Age recorded (ex) −0.47
1 = Globulin (ve) −0.46
1 = Sex (v) +0.34
0.167 = Recent utilization (fr) +0.26
0 = Acute pancreatitis (ex) −0.22
0 = Anion gap (ex) −0.21
−0.133 = Glucose (v) −0.16
0 = Propofol (ld) +0.13
0 = LVEF (%) (ve) +0.11
−0.556 = Prior utilization (fr) −0.11
0 = Pneumo Ag (ex) −0.11
0 = FHx cancer (ld) −0.11
0 = Chemo/IO visit (ld) −0.1
0 = Abd pain (ex) −0.08
0 = Urobilinogen (U) (ex) −0.07
0 = DM (no comp.) (ld) −0.07
0 = RDW (fL) (ve) −0.05
0 = WBC (corr.) (ex) −0.05
0 = RDW (ratio) (v) −0.05
0 = Albuterol (ex) −0.04
0 = Chronic pain (ex) +0.03
0 = Donepezil (ex) +0.03
0 = Pneumo vax 6B (ex) −0.03
0 = Flu A (H1N1) Ag (ex) −0.03
0 = Direct bili (ex) −0.02
0 = Urine protein (fr) +0.02
0 = Senna (ld) +0.02
0 = Joint pain (ex) +0.02
0 = MCHC (v) −0.01
0 = eGFR (v) +0.01
0 = Carvedilol (ex) +0.01
0 = Vasc headache (ld) −0.01
20 other features +0
−16 −15 −14 −13 −12
E[f(X)] = −12.348

(a)

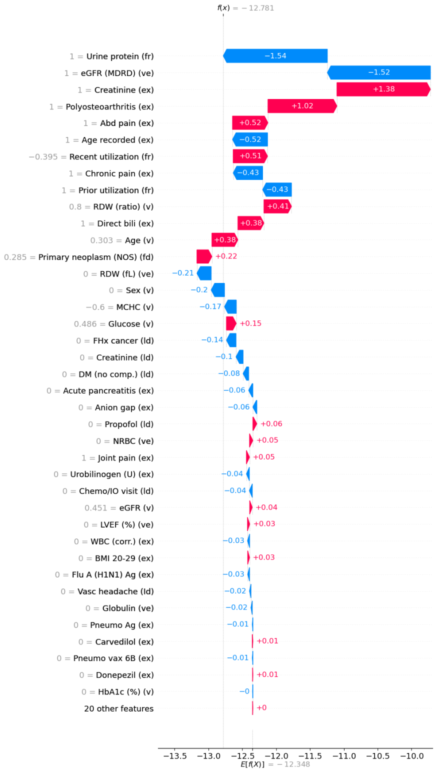
f(x) = −12.781
1 = Urine protein (fr) −1.54
1 = eGFR (MDRD) (ve) −1.52
1 = Creatinine (ex) +1.38
1 = Polyosteoarthritis (ex) +1.02
1 = Abd pain (ex) +0.52
1 = Age recorded (ex) −0.52
−0.395 = Recent utilization (fr) +0.51
1 = Chronic pain (ex) −0.43
1 = Prior utilization (fr) −0.43
0.8 = RDW (ratio) (v) +0.41
1 = Direct bili (ex) +0.38
0.303 = Age (v) +0.38
0.285 = Primary neoplasm (NOS) (fd) +0.22
0 = RDW (fL) (ve) −0.21
0 = Sex (v) −0.2
−0.6 = MCHC (v) −0.17
0.486 = Glucose (v) +0.15
0 = FHx cancer (ld) −0.14
0 = Creatinine (ld) −0.1
0 = DM (no comp.) (ld) −0.08
0 = Acute pancreatitis (ex) −0.06
0 = Anion gap (ex) −0.06
0 = Propofol (ld) +0.06
0 = NRBC (ve) +0.05
1 = Joint pain (ex) +0.05
0 = Urobilinogen (U) (ex) −0.04
0 = Chemo/IO visit (ld) −0.04
0.451 = eGFR (v) +0.04
0 = LVEF (%) (ve) +0.03
0 = WBC (corr.) (ex) −0.03
0 = BMI 20-29 (ex) +0.03
0 = Flu A (H1N1) Ag (ex) −0.03
0 = Vasc headache (ld) −0.02
0 = Globulin (ve) −0.02
0 = Pneumo Ag (ex) −0.01
0 = Carvedilol (ex) +0.01
0 = Pneumo vax 6B (ex) −0.01
0 = Donepezil (ex) +0.01
0 = HbA1c (%) (v) −0
20 other features +0
−13.5 −13.0 −12.5 −12.0 −11.5 −11.0 −10.5 −10.0
E[f(X)] = −12.348

(b)

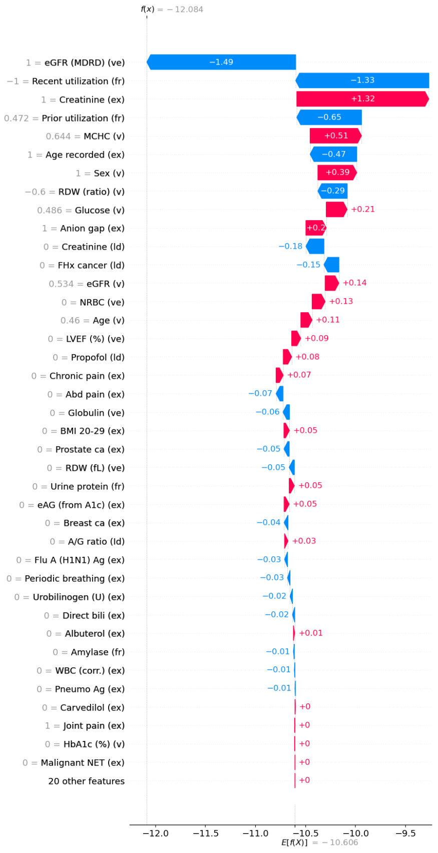
f(x) = −12.084
1 = eGFR (MDRD) (ve) −1.49
−1 = Recent utilization (fr) −1.33
1 = Creatinine (ex) +1.32
0.472 = Prior utilization (fr) −0.65
0.644 = MCHC (v) +0.51
1 = Age recorded (ex) −0.47
1 = Sex (v) +0.39
−0.6 = RDW (ratio) (v) −0.29
0.486 = Glucose (v) +0.21
1 = Anion gap (ex) +0.2
0 = Creatinine (ld) −0.18
0 = FHx cancer (ld) −0.15
0.534 = eGFR (v) +0.14
0 = NRBC (ve) +0.13
0.46 = Age (v) +0.11
0 = LVEF (%) (ve) +0.09
0 = Propofol (ld) +0.08
0 = Chronic pain (ex) +0.07
0 = Abd pain (ex) −0.07
0 = Globulin (ve) −0.06
0 = BMI 20-29 (ex) +0.05
0 = Prostate ca (ex) −0.05
0 = RDW (fL) (ve) −0.05
0 = Urine protein (fr) +0.05
0 = eAG (from A1c) (ex) +0.05
0 = Breast ca (ex) −0.04
0 = A/G ratio (ld) +0.03
0 = Flu A (H1N1) Ag (ex) −0.03
0 = Periodic breathing (ex) −0.03
0 = Urobilinogen (U) (ex) −0.02
0 = Direct bili (ex) −0.02
0 = Albuterol (ex) +0.01
0 = Amylase (fr) −0.01
0 = WBC (corr.) (ex) −0.01
0 = Pneumo Ag (ex) −0.01
0 = Carvedilol (ex) +0
1 = Joint pain (ex) +0
0 = HbA1c (%) (v) +0
0 = Malignant NET (ex) +0
20 other features +0
−12.0 −11.5 −11.0 −10.5 −10.0 −9.5
E[f(X)] = −10.606

(c)

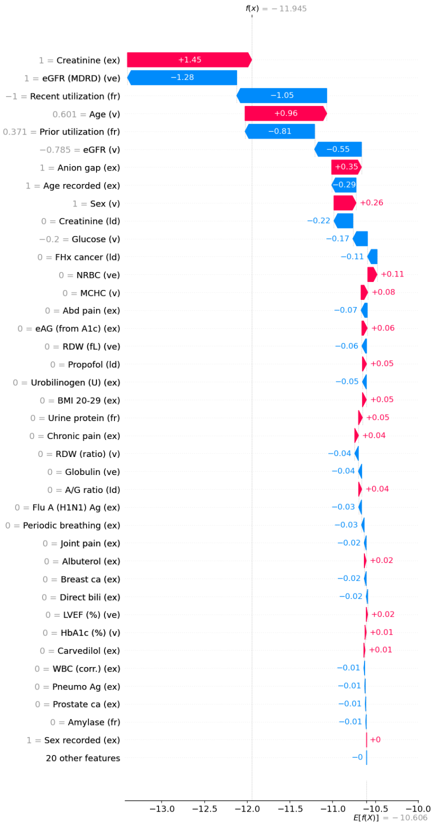
f(x) = −11.945
1 = Creatinine (ex) +1.45
1 = eGFR (MDRD) (ve) −1.28
−1 = Recent utilization (fr) −1.05
0.601 = Age (v) +0.96
0.371 = Prior utilization (fr) −0.81
−0.785 = eGFR (v) −0.55
1 = Anion gap (ex) +0.35
1 = Age recorded (ex) −0.29
1 = Sex (v) +0.26
0 = Creatinine (ld) −0.22
−0.2 = Glucose (v) −0.17
0 = FHx cancer (ld) −0.11
0 = NRBC (ve) +0.11
0 = MCHC (v) +0.08
0 = Abd pain (ex) −0.07
0 = eAG (from A1c) (ex) +0.06
0 = RDW (fL) (ve) −0.06
0 = Propofol (ld) +0.05
0 = Urobilinogen (U) (ex) −0.05
0 = BMI 20-29 (ex) +0.05
0 = Urine protein (fr) +0.05
0 = Chronic pain (ex) +0.04
0 = RDW (ratio) (v) −0.04
0 = Globulin (ve) −0.04
0 = A/G ratio (ld) +0.04
0 = Flu A (H1N1) Ag (ex) −0.03
0 = Periodic breathing (ex) −0.03
0 = Joint pain (ex) −0.02
0 = Albuterol (ex) +0.02
0 = Breast ca (ex) −0.02
0 = Direct bili (ex) −0.02
0 = LVEF (%) (ve) +0.02
0 = HbA1c (%) (v) +0.01
0 = Carvedilol (ex) +0.01
0 = WBC (corr.) (ex) −0.01
0 = Pneumo Ag (ex) −0.01
0 = Prostate ca (ex) −0.01
0 = Amylase (fr) −0.01
1 = Sex recorded (ex) +0
20 other features −0
−13.0 −12.5 −12.0 −11.5 −11.0 −10.5 −10.0
E[f(X)] = −10.606

(d)

Figure S3: **SHAP feature contributions for PRISM at SIR 5 and SIR 16**: **(a-b)** Waterfall plots for two randomly selected patients from the SIR 5 risk group. **(c-d)** Waterfall plots for two randomly selected patients from the SIR 16 risk group. Red arrows increase the SHAP value and therefore risk score relative to the expectation for a population, blue arrows decrease the SHAP value and therefore risk score relative to the expectation for a population. Longer arrows = stronger effect. The feature value is in gray to the right of the feature label. Beeswarm plots and an additional randomly selected patient's waterfall plot for each risk group can be found in Figure 4.

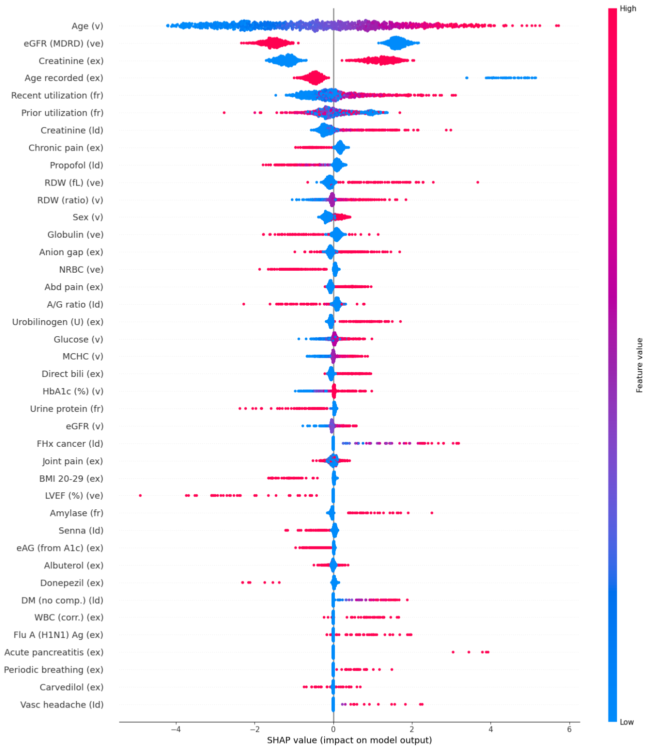

Age (v)
eGFR (MDRD) (ve)
Creatinine (ex)
Age recorded (ex)
Recent utilization (fr)
Prior utilization (fr)
Creatinine (ld)
Chronic pain (ex)
Propofol (ld)
RDW (fL) (ve)
RDW (ratio) (v)
Sex (v)
Globulin (ve)
Anion gap (ex)
NRBC (ve)
Abd pain (ex)
A/G ratio (ld)
Urobilinogen (U) (ex)
Glucose (v)
MCHC (v)
Direct bili (ex)
HbA1c (%) (v)
Urine protein (fr)
eGFR (v)
FHx cancer (ld)
Joint pain (ex)
BMI 20-29 (ex)
LVEF (%) (ve)
Amylase (fr)
Senna (ld)
eAG (from A1c) (ex)
Albuterol (ex)
Donepezil (ex)
DM (no comp.) (ld)
WBC (corr.) (ex)
Flu A (H1N1) Ag (ex)
Acute pancreatitis (ex)
Periodic breathing (ex)
Carvedilol (ex)
Vasc headache (ld)
−4
−2
0
2
4
6
SHAP value (impact on model output)
High
Feature value
Low


(a)

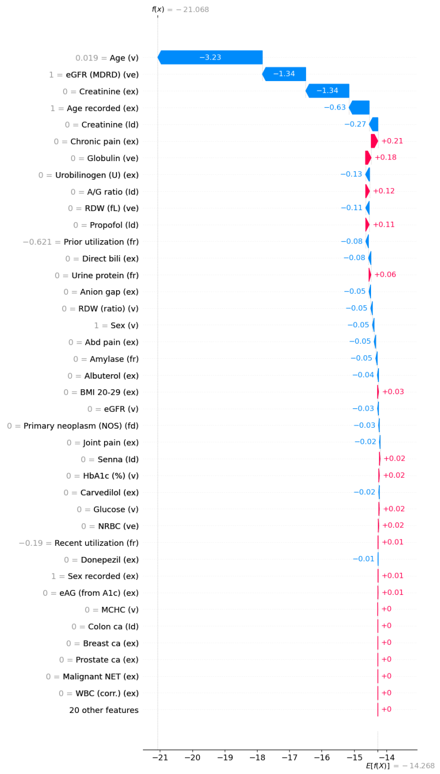

f(x) = −21.068
0.019 = Age (v) −3.23
1 = eGFR (MDRD) (ve) −1.34
0 = Creatinine (ex) −1.34
1 = Age recorded (ex) −0.63
0 = Creatinine (ld) −0.27
0 = Chronic pain (ex) +0.21
0 = Globulin (ve) +0.18
0 = Urobilinogen (U) (ex) −0.13
0 = A/G ratio (ld) +0.12
0 = RDW (fL) (ve) −0.11
0 = Propofol (ld) +0.11
−0.621 = Prior utilization (fr) −0.08
0 = Direct bili (ex) −0.08
0 = Urine protein (fr) +0.06
0 = Anion gap (ex) −0.05
0 = RDW (ratio) (v) −0.05
1 = Sex (v) −0.05
0 = Abd pain (ex) −0.05
0 = Amylase (fr) −0.05
0 = Albuterol (ex) −0.04
0 = BMI 20-29 (ex) +0.03
0 = eGFR (v) −0.03
0 = Primary neoplasm (NOS) (fd) −0.03
0 = Joint pain (ex) −0.02
0 = Senna (ld) +0.02
0 = HbA1c (%) (v) +0.02
0 = Carvedilol (ex) −0.02
0 = Glucose (v) +0.02
0 = NRBC (ve) +0.02
−0.19 = Recent utilization (fr) +0.01
0 = Donepezil (ex) −0.01
1 = Sex recorded (ex) +0.01
0 = eAG (from A1c) (ex) +0.01
0 = MCHC (v) +0
0 = Colon ca (ld) +0
0 = Breast ca (ex) +0
0 = Prostate ca (ex) +0
0 = Malignant NET (ex) +0
0 = WBC (corr.) (ex) +0
20 other features +0
−21
−20
−19
−18
−17
−16
−15
−14
E[f(X)] = −14.268


(b)

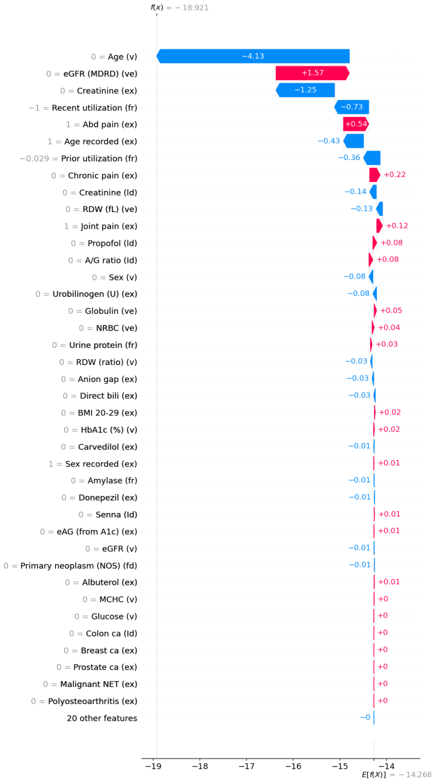

f(x) = −18.921
0 = Age (v) −4.13
0 = eGFR (MDRD) (ve) +1.57
0 = Creatinine (ex) −1.25
−1 = Recent utilization (fr) −0.73
1 = Abd pain (ex) +0.54
1 = Age recorded (ex) −0.43
−0.029 = Prior utilization (fr) −0.36
0 = Chronic pain (ex) +0.22
0 = Creatinine (ld) −0.14
0 = RDW (fL) (ve) −0.13
1 = Joint pain (ex) +0.12
0 = Propofol (ld) +0.08
0 = A/G ratio (ld) +0.08
0 = Sex (v) −0.08
0 = Urobilinogen (U) (ex) −0.08
0 = Globulin (ve) +0.05
0 = NRBC (ve) +0.04
0 = Urine protein (fr) +0.03
0 = RDW (ratio) (v) −0.03
0 = Anion gap (ex) −0.03
0 = Direct bili (ex) −0.03
0 = BMI 20-29 (ex) +0.02
0 = HbA1c (%) (v) +0.02
0 = Carvedilol (ex) −0.01
1 = Sex recorded (ex) +0.01
0 = Amylase (fr) −0.01
0 = Donepezil (ex) −0.01
0 = Senna (ld) +0.01
0 = eAG (from A1c) (ex) +0.01
0 = eGFR (v) −0.01
0 = Primary neoplasm (NOS) (fd) −0.01
0 = Albuterol (ex) +0.01
0 = MCHC (v) +0
0 = Glucose (v) +0
0 = Colon ca (ld) +0
0 = Breast ca (ex) +0
0 = Prostate ca (ex) +0
0 = Malignant NET (ex) +0
0 = Polyosteoarthritis (ex) +0
20 other features −0
−19
−18
−17
−16
−15
−14
E[f(X)] = −14.268


(c)

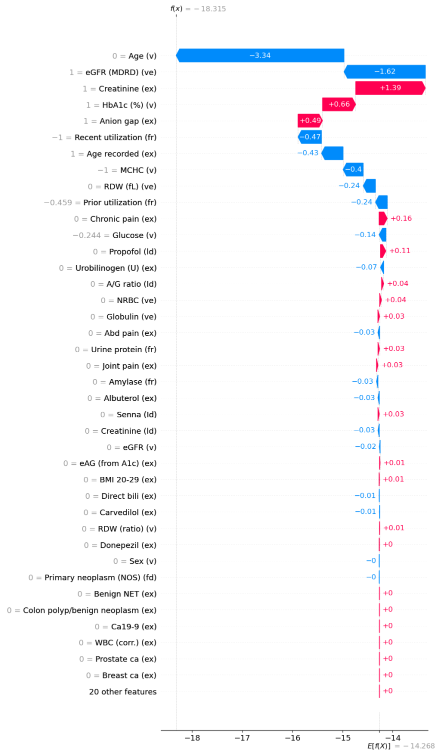

f(x) = −18.315
0 = Age (v) −3.34
1 = eGFR (MDRD) (ve) −1.62
1 = Creatinine (ex) +1.39
1 = HbA1c (%) (v) +0.66
1 = Anion gap (ex) +0.49
−1 = Recent utilization (fr) −0.47
1 = Age recorded (ex) −0.43
−1 = MCHC (v) −0.4
0 = RDW (fL) (ve) −0.24
−0.459 = Prior utilization (fr) −0.24
0 = Chronic pain (ex) +0.16
−0.244 = Glucose (v) −0.14
0 = Propofol (ld) +0.11
0 = Urobilinogen (U) (ex) −0.07
0 = A/G ratio (ld) +0.04
0 = NRBC (ve) +0.04
0 = Globulin (ve) +0.03
0 = Abd pain (ex) −0.03
0 = Urine protein (fr) +0.03
0 = Joint pain (ex) +0.03
0 = Amylase (fr) −0.03
0 = Albuterol (ex) −0.03
0 = Senna (ld) +0.03
0 = Creatinine (ld) −0.03
0 = eGFR (v) −0.02
0 = eAG (from A1c) (ex) +0.01
0 = BMI 20-29 (ex) +0.01
0 = Direct bili (ex) −0.01
0 = Carvedilol (ex) −0.01
0 = RDW (ratio) (v) +0.01
0 = Donepezil (ex) +0
0 = Sex (v) −0
0 = Primary neoplasm (NOS) (fd) −0
0 = Benign NET (ex) +0
0 = Colon polyp/benign neoplasm (ex) +0
0 = Ca19-9 (ex) +0
0 = WBC (corr.) (ex) +0
0 = Prostate ca (ex) +0
0 = Breast ca (ex) +0
20 other features +0
−18
−17
−16
−15
−14
E[f(X)] = −14.268


(d)

Figure S4: **SHAP feature contributions for PRISM at SIR 3.5**: **(a)** Beeswarm plot showing the distribution and direction of feature contributions among patients flagged at SIR 3.5. Each point represents a patient-feature pair; color indicates the feature value (red = higher values, blue = lower values), and horizontal position reflects the SHAP value (contribution to the predicted PDAC risk). Features are ordered by risk group mean SHAP value, highlighting the dominant predictors that characterize each risk stratum. **(b-d)** Waterfall plots for three randomly selected patients from the SIR 3.5 risk group. Red arrows increase the SHAP value and therefore risk score relative to the expectation for a population, blue arrows decrease the SHAP value and therefore risk score relative to the expectation for a population. Longer arrows = stronger effect. The feature value is in gray to the right of the feature label.

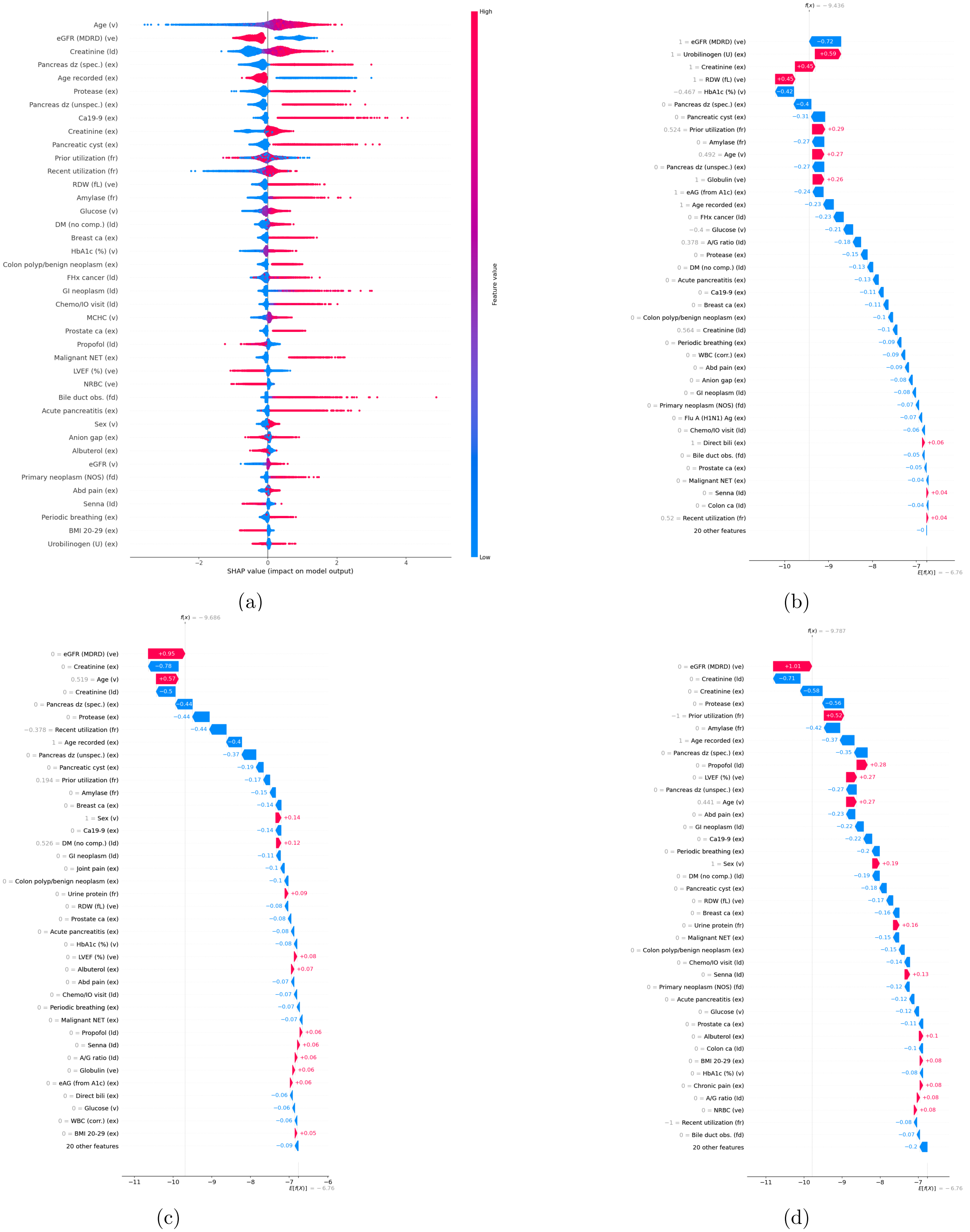
Age (v)
eGFR (MDRD) (ve)
Creatinine (ld)
Pancreas dz (spec.) (ex)
Age recorded (ex)
Protease (ex)
Pancreas dz (unspec.) (ex)
Ca19-9 (ex)
Creatinine (ex)
Pancreatic cyst (ex)
Prior utilization (fr)
Recent utilization (fr)
RDW (fL) (ve)
Amylase (fr)
Glucose (v)
DM (no comp.) (ld)
Breast ca (ex)
HbA1c (%) (v)
Colon polyp/benign neoplasm (ex)
FHx cancer (ld)
GI neoplasm (ld)
Chemo/IO visit (ld)
MCHC (v)
Prostate ca (ex)
Propofol (ld)
Malignant NET (ex)
LVEF (%) (ve)
NRBC (ve)
Bile duct obs. (fd)
Acute pancreatitis (ex)
Sex (v)
Anion gap (ex)
Albuterol (ex)
eGFR (v)
Primary neoplasm (NOS) (fd)
Abd pain (ex)
Senna (ld)
Periodic breathing (ex)
BMI 20-29 (ex)
Urobilinogen (U) (ex)
−2
0
2
4
SHAP value (impact on model output)
High
Feature value
Low
(a)
f(x) = −9.436
1 = eGFR (MDRD) (ve) −0.72
1 = Urobilinogen (U) (ex) +0.59
1 = Creatinine (ex) +0.45
1 = RDW (fL) (ve) +0.45
−0.467 = HbA1c (%) (v) −0.42
0 = Pancreas dz (spec.) (ex) −0.4
0 = Pancreatic cyst (ex) −0.31
0.524 = Prior utilization (fr) +0.29
0 = Amylase (fr) −0.27
0.492 = Age (v) +0.27
0 = Pancreas dz (unspec.) (ex) −0.27
1 = Globulin (ve) +0.26
1 = eAG (from A1c) (ex) −0.24
1 = Age recorded (ex) −0.23
0 = FHx cancer (ld) −0.23
−0.4 = Glucose (v) −0.21
0.378 = A/G ratio (ld) −0.18
0 = Protease (ex) −0.15
0 = DM (no comp.) (ld) −0.13
0 = Acute pancreatitis (ex) −0.13
0 = Ca19-9 (ex) −0.11
0 = Breast ca (ex) −0.11
0 = Colon polyp/benign neoplasm (ex) −0.1
0.564 = Creatinine (ld) −0.1
0 = Periodic breathing (ex) −0.09
0 = WBC (corr.) (ex) −0.09
0 = Abd pain (ex) −0.09
0 = Anion gap (ex) −0.08
0 = GI neoplasm (ld) −0.08
0 = Primary neoplasm (NOS) (fd) −0.07
0 = Flu A (H1N1) Ag (ex) −0.07
0 = Chemo/IO visit (ld) −0.06
1 = Direct bili (ex) +0.06
0 = Bile duct obs. (fd) −0.05
0 = Prostate ca (ex) −0.05
0 = Malignant NET (ex) −0.04
0 = Senna (ld) +0.04
0 = Colon ca (ld) −0.04
0.52 = Recent utilization (fr) +0.04
20 other features −0
−10
−9
−8
−7
E[f(X)] = −6.76
(b)
f(x) = −9.686
0 = eGFR (MDRD) (ve) +0.95
0 = Creatinine (ex) −0.78
0.519 = Age (v) +0.57
0 = Creatinine (ld) −0.5
0 = Pancreas dz (spec.) (ex) −0.44
0 = Protease (ex) −0.44
−0.378 = Recent utilization (fr) −0.44
1 = Age recorded (ex) −0.4
0 = Pancreas dz (unspec.) (ex) −0.37
0 = Pancreatic cyst (ex) −0.19
0.194 = Prior utilization (fr) −0.17
0 = Amylase (fr) −0.15
0 = Breast ca (ex) −0.14
1 = Sex (v) +0.14
0 = Ca19-9 (ex) −0.14
0.526 = DM (no comp.) (ld) +0.12
0 = GI neoplasm (ld) −0.11
0 = Joint pain (ex) −0.1
0 = Colon polyp/benign neoplasm (ex) −0.1
0 = Urine protein (fr) +0.09
0 = RDW (fL) (ve) −0.08
0 = Prostate ca (ex) −0.08
0 = Acute pancreatitis (ex) −0.08
0 = HbA1c (%) (v) −0.08
0 = LVEF (%) (ve) +0.08
0 = Albuterol (ex) +0.07
0 = Abd pain (ex) −0.07
0 = Chemo/IO visit (ld) −0.07
0 = Periodic breathing (ex) −0.07
0 = Malignant NET (ex) −0.07
0 = Propofol (ld) +0.06
0 = Senna (ld) +0.06
0 = A/G ratio (ld) +0.06
0 = Globulin (ve) +0.06
0 = eAG (from A1c) (ex) +0.06
0 = Direct bili (ex) −0.06
0 = Glucose (v) −0.06
0 = WBC (corr.) (ex) −0.06
0 = BMI 20-29 (ex) +0.05
20 other features −0.09
−11
−10
−9
−8
−7
−6
E[f(X)] = −6.76
(c)
f(x) = −9.787
0 = eGFR (MDRD) (ve) +1.01
0 = Creatinine (ld) −0.71
0 = Creatinine (ex) −0.58
0 = Protease (ex) −0.56
−1 = Prior utilization (fr) +0.52
0 = Amylase (fr) −0.42
1 = Age recorded (ex) −0.37
0 = Pancreas dz (spec.) (ex) −0.35
0 = Propofol (ld) +0.28
0 = LVEF (%) (ve) +0.27
0 = Pancreas dz (unspec.) (ex) −0.27
0.441 = Age (v) +0.27
0 = Abd pain (ex) −0.23
0 = GI neoplasm (ld) −0.22
0 = Ca19-9 (ex) −0.22
0 = Periodic breathing (ex) −0.2
1 = Sex (v) +0.19
0 = DM (no comp.) (ld) −0.19
0 = Pancreatic cyst (ex) −0.18
0 = RDW (fL) (ve) −0.17
0 = Breast ca (ex) −0.16
0 = Urine protein (fr) +0.16
0 = Malignant NET (ex) −0.15
0 = Colon polyp/benign neoplasm (ex) −0.15
0 = Chemo/IO visit (ld) −0.14
0 = Senna (ld) +0.13
0 = Primary neoplasm (NOS) (fd) −0.12
0 = Acute pancreatitis (ex) −0.12
0 = Glucose (v) −0.12
0 = Prostate ca (ex) −0.11
0 = Albuterol (ex) +0.1
0 = Colon ca (ld) −0.1
0 = BMI 20-29 (ex) +0.08
0 = HbA1c (%) (v) −0.08
0 = Chronic pain (ex) +0.08
0 = A/G ratio (ld) +0.08
0 = NRBC (ve) +0.08
−1 = Recent utilization (fr) −0.08
0 = Bile duct obs. (fd) −0.07
20 other features −0.2
−11
−10
−9
−8
−7
E[f(X)] = −6.76
(d)

Figure S5: **SHAP feature contributions for PRISM at SIR 30**: **(a)** Beeswarm plot showing the distribution and direction of feature contributions among patients flagged at SIR 31. Each point represents a patient-feature pair; color indicates the feature value (red = higher values, blue = lower values), and horizontal position reflects the SHAP value (contribution to the predicted PDAC risk). Features are ordered by risk group mean SHAP value, highlighting the dominant predictors that characterize each risk stratum. **(b-d)** Waterfall plots for three randomly selected patients from the SIR 31 risk group. Red arrows increase the SHAP value and therefore risk score relative to the expectation for a population, blue arrows decrease the SHAP value and therefore risk score relative to the expectation for a population. Longer arrows = stronger effect. The feature value is in gray to the right of the feature label.

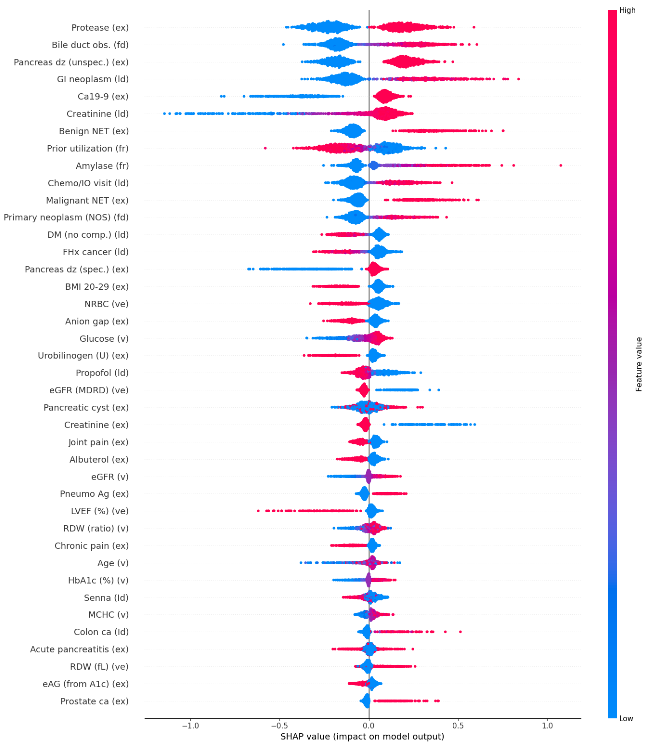

Protease (ex)
Bile duct obs. (fd)
Pancreas dz (unspec.) (ex)
GI neoplasm (ld)
Ca19-9 (ex)
Creatinine (ld)
Benign NET (ex)
Prior utilization (fr)
Amylase (fr)
Chemo/IO visit (ld)
Malignant NET (ex)
Primary neoplasm (NOS) (fd)
DM (no comp.) (ld)
FHx cancer (ld)
Pancreas dz (spec.) (ex)
BMI 20-29 (ex)
NRBC (ve)
Anion gap (ex)
Glucose (v)
Urobilinogen (U) (ex)
Propofol (ld)
eGFR (MDRD) (ve)
Pancreatic cyst (ex)
Creatinine (ex)
Joint pain (ex)
Albuterol (ex)
eGFR (v)
Pneumo Ag (ex)
LVEF (%) (ve)
RDW (ratio) (v)
Chronic pain (ex)
Age (v)
HbA1c (%) (v)
Senna (ld)
MCHC (v)
Colon ca (ld)
Acute pancreatitis (ex)
RDW (fL) (ve)
eAG (from A1c) (ex)
Prostate ca (ex)
−1.0 −0.5 0.0 0.5 1.0
SHAP value (impact on model output)
High
Feature value
Low


(a)

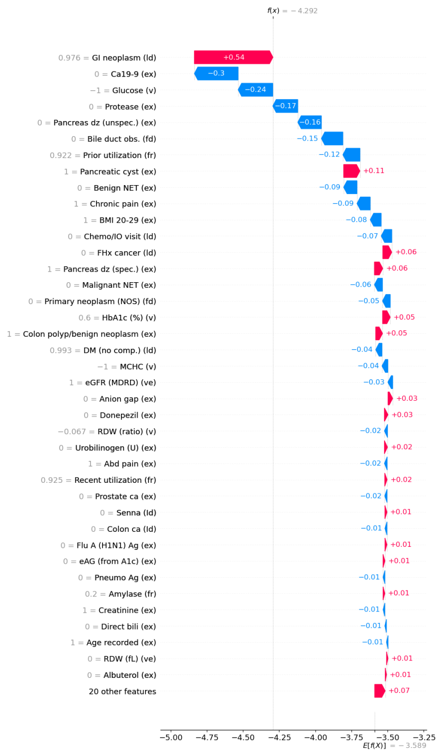

f(x) = −4.292
0.976 = GI neoplasm (ld) +0.54
0 = Ca19-9 (ex) −0.3
−1 = Glucose (v) −0.24
0 = Protease (ex) −0.17
0 = Pancreas dz (unspec.) (ex) −0.16
0 = Bile duct obs. (fd) −0.15
0.922 = Prior utilization (fr) −0.12
1 = Pancreatic cyst (ex) +0.11
0 = Benign NET (ex) −0.09
1 = Chronic pain (ex) −0.09
1 = BMI 20-29 (ex) −0.08
0 = Chemo/IO visit (ld) −0.07
0 = FHx cancer (ld) +0.06
1 = Pancreas dz (spec.) (ex) +0.06
0 = Malignant NET (ex) −0.06
0 = Primary neoplasm (NOS) (fd) −0.05
0.6 = HbA1c (%) (v) +0.05
1 = Colon polyp/benign neoplasm (ex) +0.05
0.993 = DM (no comp.) (ld) −0.04
−1 = MCHC (v) −0.04
1 = eGFR (MDRD) (ve) −0.03
0 = Anion gap (ex) +0.03
0 = Donepezil (ex) +0.03
−0.067 = RDW (ratio) (v) −0.02
0 = Urobilinogen (U) (ex) +0.02
1 = Abd pain (ex) −0.02
0.925 = Recent utilization (fr) +0.02
0 = Prostate ca (ex) −0.02
0 = Senna (ld) +0.01
0 = Colon ca (ld) −0.01
0 = Flu A (H1N1) Ag (ex) +0.01
0 = eAG (from A1c) (ex) +0.01
0 = Pneumo Ag (ex) −0.01
0.2 = Amylase (fr) +0.01
1 = Creatinine (ex) −0.01
0 = Direct bili (ex) −0.01
1 = Age recorded (ex) −0.01
0 = RDW (fL) (ve) +0.01
0 = Albuterol (ex) +0.01
20 other features +0.07
−5.00 −4.75 −4.50 −4.25 −4.00 −3.75 −3.50 −3.25
E[f(X)] = −3.589


(b)

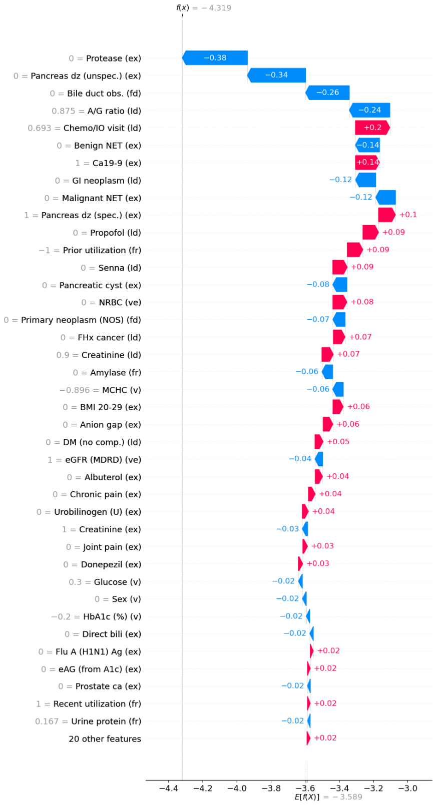

f(x) = −4.319
0 = Protease (ex) −0.38
0 = Pancreas dz (unspec.) (ex) −0.34
0 = Bile duct obs. (fd) −0.26
0.875 = A/G ratio (ld) −0.24
0.693 = Chemo/IO visit (ld) +0.2
0 = Benign NET (ex) −0.14
1 = Ca19-9 (ex) +0.14
0 = GI neoplasm (ld) −0.12
0 = Malignant NET (ex) −0.12
1 = Pancreas dz (spec.) (ex) +0.1
0 = Propofol (ld) +0.09
−1 = Prior utilization (fr) +0.09
0 = Senna (ld) +0.09
0 = Pancreatic cyst (ex) −0.08
0 = NRBC (ve) +0.08
0 = Primary neoplasm (NOS) (fd) −0.07
0 = FHx cancer (ld) +0.07
0.9 = Creatinine (ld) +0.07
0 = Amylase (fr) −0.06
−0.896 = MCHC (v) −0.06
0 = BMI 20-29 (ex) +0.06
0 = Anion gap (ex) +0.06
0 = DM (no comp.) (ld) +0.05
1 = eGFR (MDRD) (ve) −0.04
0 = Albuterol (ex) +0.04
0 = Chronic pain (ex) +0.04
0 = Urobilinogen (U) (ex) +0.04
1 = Creatinine (ex) −0.03
0 = Joint pain (ex) +0.03
0 = Donepezil (ex) +0.03
0.3 = Glucose (v) −0.02
0 = Sex (v) −0.02
−0.2 = HbA1c (%) (v) −0.02
0 = Direct bili (ex) −0.02
0 = Flu A (H1N1) Ag (ex) +0.02
0 = eAG (from A1c) (ex) +0.02
0 = Prostate ca (ex) −0.02
1 = Recent utilization (fr) +0.02
0.167 = Urine protein (fr) −0.02
20 other features +0.02
−4.4 −4.2 −4.0 −3.8 −3.6 −3.4 −3.2 −3.0
E[f(X)] = −3.589


(c)

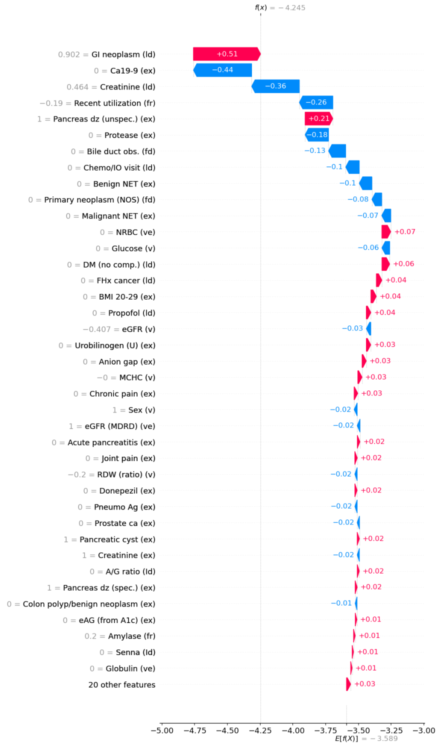

f(x) = −4.245
0.902 = GI neoplasm (ld) +0.51
0 = Ca19-9 (ex) −0.44
0.464 = Creatinine (ld) −0.36
−0.19 = Recent utilization (fr) −0.26
1 = Pancreas dz (unspec.) (ex) +0.21
0 = Protease (ex) −0.18
0 = Bile duct obs. (fd) −0.13
0 = Chemo/IO visit (ld) −0.1
0 = Benign NET (ex) −0.1
0 = Primary neoplasm (NOS) (fd) −0.08
0 = Malignant NET (ex) −0.07
0 = NRBC (ve) +0.07
0 = Glucose (v) −0.06
0 = DM (no comp.) (ld) +0.06
0 = FHx cancer (ld) +0.04
0 = BMI 20-29 (ex) +0.04
0 = Propofol (ld) +0.04
−0.407 = eGFR (v) −0.03
0 = Urobilinogen (U) (ex) +0.03
0 = Anion gap (ex) +0.03
−0 = MCHC (v) +0.03
0 = Chronic pain (ex) +0.03
1 = Sex (v) −0.02
1 = eGFR (MDRD) (ve) −0.02
0 = Acute pancreatitis (ex) +0.02
0 = Joint pain (ex) +0.02
−0.2 = RDW (ratio) (v) −0.02
0 = Donepezil (ex) +0.02
0 = Pneumo Ag (ex) −0.02
0 = Prostate ca (ex) −0.02
1 = Pancreatic cyst (ex) +0.02
1 = Creatinine (ex) −0.02
0 = A/G ratio (ld) +0.02
1 = Pancreas dz (spec.) (ex) +0.02
0 = Colon polyp/benign neoplasm (ex) −0.01
0 = eAG (from A1c) (ex) +0.01
0.2 = Amylase (fr) +0.01
0 = Senna (ld) +0.01
0 = Globulin (ve) +0.01
20 other features +0.03
−5.00 −4.75 −4.50 −4.25 −4.00 −3.75 −3.50 −3.25 −3.00
E[f(X)] = −3.589


(d)

Figure S6: **SHAP feature contributions for PRISM greater than SIR 31**: **(a)** Beeswarm plot showing the distribution and direction of feature contributions among patients flagged at SIR greater than 31. Each point represents a patient-feature pair; color indicates the feature value (red = higher values, blue = lower values), and horizontal position reflects the SHAP value (contribution to the predicted PDAC risk). Features are ordered by risk group mean SHAP value, highlighting the dominant predictors that characterize each risk stratum. **(b-d)** Waterfall plots for three randomly selected patients from the greater than SIR 31 risk group. Red arrows increase the SHAP value and therefore risk score relative to the expectation for a population, blue arrows decrease the SHAP value and therefore risk score relative to the expectation for a population. Longer arrows = stronger effect. The feature value is in gray to the right of the feature label.

Table S2: **Prespecified operating points and decision metrics at 1-30 months.** PRISM was deployed in April 2023 in a silent, non-interventional manner across 44 TriNetX health care organizations. Performance is shown at prespecified PRISM operating points carried forward from retrospective derivation to represent fixed-specificity thresholds (85%–99.9%). The raw PRISM score cutoff for each threshold is reported as "Cutoff." "Flagged(%)" reports the number and percentage of patients flagged in the overall population. For each operating point, sensitivity reflects incident PDAC within the past months of the study; PPV reflects the observed cumulative incidence among flagged patients; NNS is the reciprocal of PPV; and SIR compares observed incidence in the flagged group with expected incidence based on background rates. Timing metrics summarize time from first high-risk threshold crossing ("first high-risk flag") to diagnosis (MEPD HiRisk) and time from deployment to diagnosis (MEPD Deploy). Values in parentheses are 95% CIs. Prespecified cutoffs do not represent observed 30-month absolute risk.

| **Cutoff** | **Flagged(%)** | **Sens.** | **Spec.** | **PPV** | **NNS** | **SIR** | **MEPD HiRisk(d)** | **MEPD Deploy(d)** |
|---|---|---|---|---|---|---|---|---|
| AUC at 0m 0.777 (0.747,0.807), risk=0.004468% | | | | | | | | |
| 7.8022 | 758,799 (15.00) | 53.10% | 85.00% | 0.02% | 6323.3 | 3.5 | 1434 (1192-1872) | 13 (11-15) |
| 7.2938 | 556,500 (11.00) | 45.58% | 89.00% | 0.02% | 5402.9 | 4.1 | 1246 (926-1701) | 13 (11-17) |
| 6.6796 | 252,969 (5.00) | 33.19% | 95.00% | 0.03% | 3372.9 | 6.6 | 1037 (754-1430) | 12 (11-17) |
| 6.4058 | 151,806 (3.00) | 27.43% | 97.00% | 0.04% | 2448.5 | 9.1 | 1018 (665-1484) | 11 (10-17) |
| 5.4374 | 25,376 (0.50) | 12.39% | 99.50% | 0.11% | 906.3 | 24.7 | 845 (653-1635) | 10 (5-15) |
| 4.6214 | 5,286 (0.10) | 4.87% | 99.90% | 0.21% | 480.5 | 46.6 | 756 (275-1474) | 13(5-18) |
| AUC at 1m 0.747 (0.723,0.771), risk=0.007888% | | | | | | | | |
| 7.8023 | 758,840 (15.00) | 47.37% | 85.00% | 0.02% | 4015.0 | 3.2 | 1590 (1391-1873) | 22 (19-26) |
| 7.2938 | 556,509 (11.00) | 41.35% | 89.00% | 0.03% | 3372.8 | 3.8 | 1390 (1062-1763) | 24 (19-27) |
| 6.6796 | 253,003 (5.00) | 29.57% | 95.00% | 0.05% | 2144.1 | 5.9 | 1297 (1045-1580) | 24 (18-27) |
| 6.4059 | 151,836 (3.00) | 24.06% | 97.00% | 0.06% | 1581.6 | 8.0 | 1303 (831-1693) | 22 (16-27) |
| 5.4374 | 25,376 (0.50) | 11.03% | 99.50% | 0.17% | 576.7 | 22.0 | 1042 (653-1906) | 20 (12-29) |
| 4.6214 | 5,286 (0.10) | 4.01% | 99.90% | 0.30% | 330.4 | 38.4 | 974 (503-1574) | 18 (11-34) |
| AUC at 2m 0.742 (0.721,0.762), risk=0.01162% | | | | | | | | |
| 7.8025 | 758,914 (15.00) | 47.79% | 85.00% | 0.04% | 2700.8 | 3.2 | 1603 (1412-1859) | 41 (32-47) |
| 7.2940 | 556,576 (11.00) | 41.67% | 89.00% | 0.04% | 2271.7 | 3.8 | 1422 (1196-1727) | 41 (32-47) |
| 6.6797 | 253,059 (5.00) | 30.10% | 95.00% | 0.07% | 1429.7 | 6.0 | 1278 (1062-1518) | 42 (30-51) |
| 6.4060 | 151,896 (3.00) | 24.32% | 97.00% | 0.09% | 1062.2 | 8.1 | 1303 (1012-1554) | 41 (28-51) |
| 5.4374 | 25,376 (0.50) | 12.24% | 99.50% | 0.28% | 352.4 | 24.4 | 845 (625-1402) | 47 (30-60) |
| 4.6214 | 5,286 (0.10) | 4.25% | 99.90% | 0.47% | 211.4 | 40.7 | 812 (544-1346) | 38 (18-62) |
| AUC at 3m 0.744 (0.726,0.761), risk=0.01534% | | | | | | | | |
| 7.8026 | 758,968 (15.01) | 47.16% | 85.00% | 0.05% | 2073.7 | 3.1 | 1834 (1597-1967) | 56 (51-64) |
| 7.2942 | 556,634 (11.00) | 41.49% | 89.00% | 0.06% | 1728.7 | 3.8 | 1700 (1399-1891) | 60 (50-64) |
| 6.6798 | 253,091 (5.00) | 29.12% | 95.00% | 0.09% | 1119.9 | 5.8 | 1486 (1226-1663) | 58 (48-64) |
| 6.4060 | 151,896 (3.00) | 22.68% | 97.00% | 0.12% | 863.0 | 7.6 | 1386 (1037-1621) | 55 (44-63) |
| 5.4374 | 25,376 (0.50) | 11.08% | 99.50% | 0.34% | 295.1 | 22.1 | 1076 (710-1462) | 60 (42-70) |
| 4.6214 | 5,286 (0.10) | 3.87% | 99.90% | 0.57% | 176.2 | 37.0 | 1136 (657-1512) | 48 (25-71) |
| AUC at 4m 0.738 (0.722,0.754), risk=0.01888% | | | | | | | | |
| 7.8027 | 759,009 (15.01) | 46.28% | 85.00% | 0.06% | 1717.2 | 3.1 | 1826 (1603-1941) | 70 (63-77) |
| 7.2943 | 556,667 (11.01) | 40.21% | 89.00% | 0.07% | 1449.7 | 3.7 | 1695 (1456-1879) | 70 (62-76) |
| 6.6799 | 253,119 (5.00) | 27.54% | 95.00% | 0.10% | 962.4 | 5.5 | 1502 (1299-1677) | 67 (60-75) |

| | | | | | | | | |
|---|---|---|---|---|---|---|---|---|
| 6.4061 | 151,915 (3.00) | 21.05% | 97.00% | 0.13% | 755.8 | 7.0 | 1404 (1092-1648) | 62 (54-73) |
| 5.4377 | 25,398 (0.50) | 10.16% | 99.50% | 0.38% | 261.8 | 20.2 | 1076 (747-1450) | 68 (50-75) |
| 4.6214 | 5,286 (0.10) | 3.87% | 99.90% | 0.70% | 142.9 | 37.1 | 1079 (694-1512) | 69 (44-84) |
| AUC at 5m 0.739 (0.724,0.754), risk=0.02232% | | | | | | | | |
| 7.8028 | 759,065 (15.01) | 46.06% | 85.00% | 0.07% | 1459.7 | 3.1 | 1842 (1639-2018) | 83 (76-91) |
| 7.2944 | 556,719 (11.01) | 40.12% | 89.00% | 0.08% | 1229.0 | 3.6 | 1704 (1518-1899) | 83 (75-91) |
| 6.6800 | 253,163 (5.01) | 27.28% | 95.00% | 0.12% | 822.0 | 5.5 | 1520 (1331-1677) | 79 (69-89) |
| 6.4062 | 151,948 (3.00) | 20.81% | 97.00% | 0.15% | 646.6 | 6.9 | 1423 (1112-1624) | 75 (65-84) |
| 5.4377 | 25,398 (0.50) | 9.65% | 99.50% | 0.43% | 233.0 | 19.2 | 1040 (722-1426) | 73 (61-83) |
| 4.6214 | 5,286 (0.10) | 3.72% | 99.90% | 0.79% | 125.9 | 35.6 | 1082 (735-1469) | 77 (48-101) |
| AUC at 6m 0.741 (0.728,0.755), risk=0.02574% | | | | | | | | |
| 7.8029 | 759,114 (15.01) | 46.01% | 85.00% | 0.08% | 1267.3 | 3.1 | 1881 (1705-2065) | 96 (90-107) |
| 7.2946 | 556,771 (11.01) | 40.32% | 89.00% | 0.09% | 1060.5 | 3.7 | 1757 (1589-1930) | 100 (90-109) |
| 6.6800 | 253,204 (5.01) | 27.34% | 95.00% | 0.14% | 711.2 | 5.5 | 1524 (1353-1678) | 92 (83-103) |
| 6.4063 | 151,973 (3.00) | 20.74% | 97.00% | 0.18% | 562.9 | 6.9 | 1442 (1165-1648) | 87 (77-101) |
| 5.4405 | 25,543 (0.50) | 9.29% | 99.50% | 0.47% | 211.1 | 18.4 | 1073 (736-1403) | 79 (70-90) |
| 4.6214 | 5,286 (0.10) | 3.38% | 99.90% | 0.83% | 120.1 | 32.3 | 1193 (814-1512) | 82 (57-107) |
| AUC at 7m 0.738 (0.725,0.751), risk=0.02894% | | | | | | | | |
| 7.8030 | 759,157 (15.01) | 45.36% | 85.00% | 0.09% | 1143.3 | 3.0 | 1868 (1705-2036) | 111 (101-118) |
| 7.2947 | 556,818 (11.01) | 39.82% | 89.00% | 0.10% | 955.1 | 3.6 | 1782 (1614-1930) | 111 (102-119) |
| 6.6801 | 253,225 (5.01) | 26.57% | 95.00% | 0.15% | 651.0 | 5.3 | 1557 (1374-1707) | 102 (90-115) |
| 6.4065 | 152,023 (3.01) | 20.22% | 97.00% | 0.19% | 513.6 | 6.7 | 1446 (1165-1648) | 96 (84-115) |
| 5.4405 | 25,543 (0.50) | 9.08% | 99.50% | 0.52% | 192.1 | 18.0 | 1040 (736-1381) | 83 (73-105) |
| 4.6214 | 5,286 (0.10) | 3.28% | 99.90% | 0.91% | 110.1 | 31.4 | 1086 (813-1460) | 84 (62-123) |
| AUC at 8m 0.737 (0.725,0.749), risk=0.03211% | | | | | | | | |
| 7.8031 | 759,197 (15.01) | 44.89% | 85.00% | 0.10% | 1041.4 | 3.0 | 1848 (1684-2027) | 120 (113-130) |
| 7.2948 | 556,856 (11.01) | 39.41% | 89.00% | 0.11% | 870.1 | 3.6 | 1772 (1611-1907) | 120 (113-132) |
| 6.6801 | 253,245 (5.01) | 25.92% | 95.00% | 0.17% | 601.5 | 5.2 | 1532 (1374-1688) | 115 (101-125) |
| 6.4065 | 152,023 (3.01) | 19.64% | 97.00% | 0.21% | 476.6 | 6.5 | 1413 (1161-1597) | 109 (90-123) |
| 5.4405 | 25,543 (0.50) | 8.62% | 99.50% | 0.55% | 182.5 | 17.1 | 1002 (747-1274) | 90 (76-115) |
| 4.6214 | 5,286 (0.10) | 3.08% | 99.90% | 0.95% | 105.7 | 29.5 | 1112 (814-1460) | 90 (66-125) |
| AUC at 9m 0.735 (0.724,0.747), risk=0.03549% | | | | | | | | |
| 7.8032 | 759,250 (15.01) | 44.79% | 85.00% | 0.11% | 944.3 | 3.0 | 1825 (1660-1929) | 133 (125-145) |
| 7.2949 | 556,891 (11.01) | 39.00% | 89.00% | 0.13% | 795.6 | 3.5 | 1728 (1588-1865) | 133 (122-146) |
| 6.6802 | 253,279 (5.01) | 25.74% | 95.00% | 0.18% | 548.2 | 5.1 | 1514 (1353-1659) | 125 (115-141) |
| 6.4066 | 152,052 (3.01) | 19.55% | 97.00% | 0.23% | 433.2 | 6.5 | 1376 (1144-1539) | 119 (103-141) |
| 5.4405 | 25,543 (0.50) | 8.75% | 99.50% | 0.61% | 162.7 | 17.3 | 1018 (796-1216) | 109 (83-131) |
| 4.6214 | 5,286 (0.10) | 3.06% | 99.90% | 1.04% | 96.1 | 29.3 | 1065 (753-1437) | 102 (72-135) |
| AUC at 10m 0.737 (0.726,0.748), risk=0.03822% | | | | | | | | |
| 7.8033 | 759,302 (15.01) | 45.16% | 85.00% | 0.11% | 869.8 | 3.0 | 1834 (1691-1934) | 147 (135-158) |
| 7.2951 | 556,940 (11.01) | 39.11% | 89.00% | 0.14% | 736.7 | 3.6 | 1730 (1589-1865) | 147 (134-158) |
| 6.6802 | 253,305 (5.01) | 25.76% | 95.00% | 0.20% | 508.6 | 5.1 | 1518 (1374-1665) | 140 (123-154) |
| 6.4067 | 152,071 (3.01) | 19.71% | 97.00% | 0.25% | 399.1 | 6.6 | 1389 (1161-1562) | 137 (117-156) |
| 5.4405 | 25,543 (0.50) | 8.90% | 99.50% | 0.67% | 148.5 | 17.6 | 992 (796-1188) | 120 (95-156) |
| 4.6214 | 5,286 (0.10) | 3.10% | 99.90% | 1.14% | 88.1 | 29.7 | 1150 (813-1491) | 120 (82-159) |
| AUC at 11m 0.737 (0.726,0.748), risk=0.04126% | | | | | | | | |

| | | | | | | | | |
|---|---|---|---|---|---|---|---|---|
| 7.8034 | 759,340 (15.01) | 45.09% | 85.00% | 0.12% | 807.0 | 3.0 | 1840 (1716-1981) | 159 (150-172) |
| 7.2952 | 556,988 (11.01) | 39.15% | 89.00% | 0.15% | 681.7 | 3.6 | 1732 (1611-1874) | 159 (150-172) |
| 6.6803 | 253,334 (5.01) | 25.54% | 95.00% | 0.21% | 475.3 | 5.1 | 1570 (1407-1725) | 152 (135-163) |
| 6.4068 | 152,090 (3.01) | 19.45% | 97.00% | 0.27% | 374.6 | 6.5 | 1430 (1207-1597) | 151 (131-165) |
| 5.4405 | 25,543 (0.50) | 8.62% | 99.50% | 0.70% | 141.9 | 17.1 | 1028 (831-1216) | 130 (102-165) |
| 4.6214 | 5,286 (0.10) | 3.02% | 99.90% | 1.19% | 83.9 | 28.9 | 1160 (941-1491) | 124 (84-166) |
| AUC at 12m 0.738 (0.727,0.748), risk=0.04423% | | | | | | | | |
| 7.8035 | 759,384 (15.01) | 45.06% | 85.00% | 0.13% | 753.4 | 3.0 | 1894 (1788-2046) | 172 (160-186) |
| 7.2953 | 557,022 (11.01) | 39.07% | 89.00% | 0.16% | 637.3 | 3.5 | 1826 (1694-1930) | 172 (159-186) |
| 6.6804 | 253,379 (5.01) | 25.39% | 95.00% | 0.22% | 446.1 | 5.1 | 1649 (1503-1830) | 161 (149-179) |
| 6.4068 | 152,108 (3.01) | 19.31% | 97.00% | 0.28% | 352.1 | 6.4 | 1520 (1358-1666) | 160 (143-180) |
| 5.4405 | 25,543 (0.50) | 8.54% | 99.50% | 0.75% | 133.7 | 16.9 | 1052 (886-1216) | 140 (115-179) |
| 4.6214 | 5,286 (0.10) | 3.17% | 99.90% | 1.34% | 74.5 | 30.4 | 1139 (998-1461) | 139 (101-207) |
| AUC at 13m 0.737 (0.727,0.747), risk=0.04682% | | | | | | | | |
| 7.8036 | 759,422 (15.01) | 44.89% | 85.00% | 0.14% | 714.4 | 3.0 | 1921 (1818-2081) | 185 (172-199) |
| 7.2954 | 557,049 (11.01) | 38.64% | 89.00% | 0.16% | 608.8 | 3.5 | 1841 (1712-2003) | 184 (167-195) |
| 6.6804 | 253,393 (5.01) | 25.34% | 95.00% | 0.24% | 422.3 | 5.1 | 1688 (1542-1857) | 172 (157-189) |
| 6.4069 | 152,138 (3.01) | 19.34% | 97.00% | 0.30% | 332.2 | 6.4 | 1565 (1421-1719) | 172 (154-193) |
| 5.4405 | 25,543 (0.50) | 8.61% | 99.50% | 0.80% | 125.2 | 17.1 | 1102 (930-1305) | 163 (126-200) |
| 4.6214 | 5,286 (0.10) | 3.29% | 99.90% | 1.48% | 67.8 | 31.5 | 1160 (1026-1491) | 168 (120-241) |
| AUC at 14m 0.736 (0.727,0.746), risk=0.04954% | | | | | | | | |
| 7.8037 | 759,458 (15.01) | 44.69% | 85.00% | 0.15% | 678.1 | 3.0 | 1921 (1813-2072) | 197 (184-208) |
| 7.2954 | 557,080 (11.01) | 38.35% | 89.00% | 0.17% | 579.7 | 3.5 | 1844 (1722-2004) | 192 (180-203) |
| 6.6805 | 253,410 (5.01) | 24.98% | 95.00% | 0.25% | 404.8 | 5.0 | 1668 (1542-1846) | 185 (163-199) |
| 6.4070 | 152,173 (3.01) | 19.07% | 97.00% | 0.31% | 318.4 | 6.3 | 1566 (1428-1711) | 184 (162-201) |
| 5.4405 | 25,543 (0.50) | 8.54% | 99.50% | 0.84% | 119.4 | 16.9 | 1121 (950-1315) | 174 (133-212) |
| 4.6214 | 5,286 (0.10) | 3.27% | 99.90% | 1.55% | 64.5 | 31.3 | 1193 (1067-1491) | 174 (126-263) |
| AUC at 15m 0.737 (0.727,0.746), risk=0.0519% | | | | | | | | |
| 7.8038 | 759,503 (15.02) | 44.88% | 85.00% | 0.16% | 644.7 | 3.0 | 1979 (1838-2116) | 206 (194-221) |
| 7.2956 | 557,116 (11.01) | 38.40% | 89.00% | 0.18% | 552.7 | 3.5 | 1888 (1768-2037) | 201 (188-218) |
| 6.6805 | 253,427 (5.01) | 24.88% | 95.00% | 0.26% | 388.1 | 5.0 | 1728 (1593-1875) | 192 (173-208) |
| 6.4070 | 152,173 (3.01) | 18.93% | 97.00% | 0.33% | 306.2 | 6.3 | 1587 (1437-1730) | 188 (171-210) |
| 5.4405 | 25,543 (0.50) | 8.30% | 99.50% | 0.85% | 117.2 | 16.4 | 1141 (994-1363) | 180 (140-221) |
| 4.6214 | 5,286 (0.10) | 3.16% | 99.90% | 1.57% | 63.7 | 30.3 | 1212 (1081-1491) | 176 (129-263) |
| AUC at 16m 0.736 (0.727,0.746), risk=0.05417% | | | | | | | | |
| 7.8038 | 759,526 (15.02) | 44.74% | 85.00% | 0.16% | 619.5 | 3.0 | 1970 (1838-2112) | 219 (202-233) |
| 7.2956 | 557,136 (11.01) | 38.10% | 89.00% | 0.19% | 533.7 | 3.5 | 1886 (1768-2033) | 210 (197-227) |
| 6.6806 | 253,453 (5.01) | 24.64% | 95.00% | 0.27% | 375.5 | 4.9 | 1732 (1608-1866) | 199 (185-220) |
| 6.4070 | 152,173 (3.01) | 18.65% | 97.00% | 0.34% | 297.8 | 6.2 | 1590 (1442-1730) | 199 (177-221) |
| 5.4405 | 25,543 (0.50) | 8.10% | 99.50% | 0.87% | 115.1 | 16.0 | 1162 (1006-1377) | 188 (146-223) |
| 4.6214 | 5,286 (0.10) | 3.10% | 99.90% | 1.61% | 62.2 | 29.7 | 1216 (1081-1509) | 201 (129-281) |
| AUC at 17m 0.737 (0.727,0.746), risk=0.05573% | | | | | | | | |
| 7.8039 | 759,554 (15.02) | 44.63% | 85.00% | 0.17% | 603.8 | 3.0 | 1982 (1844-2125) | 222 (209-240) |
| 7.2957 | 557,156 (11.02) | 38.06% | 89.00% | 0.19% | 519.3 | 3.5 | 1894 (1797-2039) | 219 (201-234) |
| 6.6806 | 253,455 (5.01) | 24.55% | 95.00% | 0.27% | 366.3 | 4.9 | 1750 (1619-1887) | 204 (187-227) |
| 6.4071 | 152,186 (3.01) | 18.66% | 97.00% | 0.35% | 289.3 | 6.2 | 1597 (1455-1741) | 203 (185-229) |

| | | | | | | | | |
|---|---|---|---|---|---|---|---|---|
| 5.4405 | 25,543 (0.50) | 8.09% | 99.50% | 0.89% | 112.0 | 16.0 | 1206 (1054-1387) | 195 (157-234) |
| 4.6214 | 5,286 (0.10) | 3.05% | 99.90% | 1.63% | 61.5 | 29.2 | 1313 (1085-1511) | 202 (132-281) |
| AUC at 18m 0.738 (0.729,0.747), risk=0.0572% | | | | | | | | |
| 7.8040 | 759,576 (15.02) | 44.83% | 85.00% | 0.17% | 585.6 | 3.0 | 2004 (1860-2134) | 233 (219-251) |
| 7.2957 | 557,174 (11.02) | 38.13% | 89.00% | 0.20% | 505.1 | 3.5 | 1914 (1824-2058) | 224 (208-241) |
| 6.6806 | 253,493 (5.01) | 24.61% | 95.00% | 0.28% | 356.0 | 4.9 | 1768 (1636-1898) | 210 (194-233) |
| 6.4071 | 152,191 (3.01) | 18.49% | 97.00% | 0.35% | 284.5 | 6.1 | 1618 (1482-1750) | 208 (187-233) |
| 5.4405 | 25,543 (0.50) | 8.05% | 99.50% | 0.91% | 109.6 | 15.9 | 1235 (1076-1404) | 201 (163-237) |
| 4.6214 | 5,286 (0.10) | 3.08% | 99.90% | 1.68% | 59.4 | 29.4 | 1313 (1098-1511) | 214 (137-298) |
| AUC at 19m 0.738 (0.729,0.747), risk=0.05864% | | | | | | | | |
| 7.8041 | 759,616 (15.02) | 45.01% | 85.00% | 0.18% | 569.0 | 3.0 | 2002 (1853-2134) | 242 (224-258) |
| 7.2959 | 557,213 (11.02) | 38.27% | 89.00% | 0.20% | 490.9 | 3.5 | 1904 (1808-2057) | 233 (218-251) |
| 6.6806 | 253,493 (5.01) | 24.68% | 95.00% | 0.29% | 346.3 | 4.9 | 1783 (1650-1918) | 222 (200-244) |
| 6.4073 | 152,245 (3.01) | 18.51% | 97.00% | 0.36% | 277.3 | 6.1 | 1630 (1487-1758) | 216 (194-237) |
| 5.4405 | 25,543 (0.50) | 8.02% | 99.50% | 0.93% | 107.3 | 15.9 | 1245 (1086-1413) | 203 (170-249) |
| 4.6214 | 5,286 (0.10) | 3.10% | 99.90% | 1.74% | 57.5 | 29.7 | 1312 (1098-1511) | 221 (150-303) |
| AUC at 20m 0.739 (0.730,0.747), risk=0.05979% | | | | | | | | |
| 7.8041 | 759,616 (15.02) | 44.87% | 85.00% | 0.18% | 559.8 | 3.0 | 2005 (1863-2140) | 249 (230-264) |
| 7.2959 | 557,213 (11.02) | 38.19% | 89.00% | 0.21% | 482.4 | 3.5 | 1914 (1821-2058) | 237 (221-257) |
| 6.6807 | 253,501 (5.01) | 24.74% | 95.00% | 0.30% | 338.9 | 4.9 | 1805 (1657-1921) | 227 (204-254) |
| 6.4073 | 152,245 (3.01) | 18.49% | 97.00% | 0.37% | 272.4 | 6.1 | 1642 (1489-1773) | 222 (199-246) |
| 5.4405 | 25,543 (0.50) | 8.04% | 99.50% | 0.95% | 105.1 | 15.9 | 1251 (1096-1413) | 214 (173-260) |
| 4.6214 | 5,286 (0.10) | 3.14% | 99.90% | 1.80% | 55.6 | 30.1 | 1297 (1124-1509) | 237 (161-313) |
| AUC at 21m 0.739 (0.730,0.748), risk=0.06125% | | | | | | | | |
| 7.8041 | 759,638 (15.02) | 44.90% | 85.00% | 0.18% | 546.1 | 3.0 | 2025 (1881-2155) | 257 (237-271) |
| 7.2960 | 557,262 (11.02) | 38.22% | 89.00% | 0.21% | 470.7 | 3.5 | 1927 (1826-2065) | 246 (227-264) |
| 6.6807 | 253,516 (5.01) | 24.69% | 95.00% | 0.30% | 331.4 | 4.9 | 1805 (1659-1921) | 234 (210-262) |
| 6.4073 | 152,245 (3.01) | 18.53% | 97.00% | 0.38% | 265.2 | 6.2 | 1634 (1488-1766) | 230 (203-257) |
| 5.4405 | 25,543 (0.50) | 8.01% | 99.50% | 0.97% | 103.0 | 15.9 | 1251 (1096-1413) | 222 (180-264) |
| 4.6214 | 5,286 (0.10) | 3.13% | 99.90% | 1.84% | 54.5 | 30.0 | 1282 (1124-1491) | 248 (168-319) |
| AUC at 22m 0.739 (0.730,0.748), risk=0.06261% | | | | | | | | |
| 7.8042 | 759,658 (15.02) | 44.90% | 85.00% | 0.19% | 534.2 | 3.0 | 2037 (1906-2178) | 264 (247-278) |
| 7.2960 | 557,262 (11.02) | 38.24% | 89.00% | 0.22% | 460.2 | 3.5 | 1980 (1846-2077) | 253 (235-271) |
| 6.6807 | 253,531 (5.01) | 24.76% | 95.00% | 0.31% | 323.4 | 4.9 | 1834 (1685-1951) | 244 (222-268) |
| 6.4073 | 152,245 (3.01) | 18.53% | 97.00% | 0.39% | 259.4 | 6.2 | 1666 (1521-1806) | 235 (210-265) |
| 5.4405 | 25,543 (0.50) | 8.02% | 99.50% | 0.99% | 100.6 | 15.9 | 1255 (1118-1416) | 226 (188-278) |
| 4.6214 | 5,286 (0.10) | 3.09% | 99.90% | 1.85% | 53.9 | 29.6 | 1297 (1124-1509) | 252 (168-321) |
| AUC at 23m 0.739 (0.730,0.747), risk=0.06417% | | | | | | | | |
| 7.8042 | 759,684 (15.02) | 44.76% | 85.00% | 0.19% | 522.8 | 3.0 | 2059 (1924-2184) | 269 (253-285) |
| 7.2960 | 557,270 (11.02) | 38.08% | 89.00% | 0.22% | 450.9 | 3.5 | 2002 (1862-2114) | 258 (242-278) |
| 6.6808 | 253,582 (5.01) | 24.61% | 95.00% | 0.32% | 317.4 | 4.9 | 1844 (1697-1976) | 251 (227-277) |
| 6.4073 | 152,245 (3.01) | 18.36% | 97.00% | 0.39% | 255.4 | 6.1 | 1683 (1542-1815) | 240 (215-273) |
| 5.4405 | 25,543 (0.50) | 7.92% | 99.50% | 1.01% | 99.4 | 15.7 | 1262 (1122-1413) | 234 (194-278) |
| 4.6181 | 5,257 (0.10) | 3.05% | 99.90% | 1.88% | 53.1 | 29.3 | 1280 (1124-1491) | 257 (173-321) |
| AUC at 24m 0.739 (0.731,0.748), risk=0.06558% | | | | | | | | |
| 7.8043 | 759,733 (15.02) | 45.04% | 85.00% | 0.20% | 508.5 | 3.0 | 2071 (1933-2188) | 278 (263-293) |

| | | | | | | | | |
|---|---|---|---|---|---|---|---|---|
| 7.2960 | 557,292 (11.02) | 38.23% | 89.00% | 0.23% | 439.5 | 3.5 | 2004 (1865-2119) | 266 (250-286) |
| 6.6808 | 253,582 (5.01) | 24.69% | 95.00% | 0.32% | 309.6 | 4.9 | 1844 (1710-1976) | 262 (233-284) |
| 6.4074 | 152,263 (3.01) | 18.45% | 97.00% | 0.40% | 248.8 | 6.1 | 1684 (1553-1815) | 250 (223-279) |
| 5.4405 | 25,543 (0.50) | 8.02% | 99.50% | 1.04% | 96.0 | 15.9 | 1259 (1122-1392) | 243 (201-287) |
| 4.6181 | 5,257 (0.10) | 3.11% | 99.90% | 1.96% | 51.0 | 29.9 | 1276 (1098-1462) | 280 (191-348) |
| AUC at 25m 0.741 (0.732,0.749), risk=0.06698% | | | | | | | | |
| 7.8043 | 759,733 (15.02) | 45.19% | 85.00% | 0.20% | 496.2 | 3.0 | 2124 (1986-2213) | 286 (270-300) |
| 7.2961 | 557,318 (11.02) | 38.43% | 89.00% | 0.23% | 428.0 | 3.5 | 2037 (1900-2146) | 278 (258-294) |
| 6.6808 | 253,582 (5.01) | 24.76% | 95.00% | 0.33% | 302.2 | 4.9 | 1864 (1751-2022) | 269 (243-290) |
| 6.4075 | 152,273 (3.01) | 18.54% | 97.00% | 0.41% | 242.5 | 6.2 | 1720 (1597-1832) | 264 (233-288) |
| 5.4410 | 25,570 (0.51) | 8.09% | 99.50% | 1.07% | 93.3 | 16.0 | 1284 (1139-1428) | 260 (212-293) |
| 4.6181 | 5,257 (0.10) | 3.13% | 99.90% | 2.02% | 49.6 | 30.1 | 1312 (1138-1516) | 290 (202-361) |
| AUC at 26m 0.739 (0.731,0.748), risk=0.06807% | | | | | | | | |
| 7.8044 | 759,747 (15.02) | 45.08% | 85.00% | 0.20% | 489.5 | 3.0 | 2133 (2000-2242) | 289 (274-305) |
| 7.2961 | 557,327 (11.02) | 38.28% | 89.00% | 0.24% | 422.9 | 3.5 | 2039 (1903-2150) | 282 (264-299) |
| 6.6808 | 253,582 (5.01) | 24.66% | 95.00% | 0.33% | 298.7 | 4.9 | 1868 (1764-2022) | 274 (248-293) |
| 6.4075 | 152,275 (3.01) | 18.47% | 97.00% | 0.42% | 239.4 | 6.1 | 1728 (1597-1833) | 265 (235-292) |
| 5.4410 | 25,570 (0.51) | 7.99% | 99.50% | 1.08% | 93.0 | 15.8 | 1304 (1141-1428) | 264 (212-295) |
| 4.6181 | 5,257 (0.10) | 3.08% | 99.90% | 2.02% | 49.6 | 29.6 | 1318 (1150-1521) | 290 (202-361) |
| AUC at 27m 0.740 (0.732,0.748), risk=0.0692% | | | | | | | | |
| 7.8044 | 759,772 (15.02) | 45.14% | 85.00% | 0.21% | 480.9 | 3.0 | 2146 (2025-2252) | 297 (279-311) |
| 7.2962 | 557,345 (11.02) | 38.34% | 89.00% | 0.24% | 415.3 | 3.5 | 2058 (1927-2171) | 287 (267-304) |
| 6.6808 | 253,600 (5.01) | 24.69% | 95.00% | 0.34% | 293.5 | 4.9 | 1888 (1783-2030) | 279 (257-300) |
| 6.4077 | 152,348 (3.01) | 18.51% | 97.00% | 0.43% | 235.1 | 6.1 | 1744 (1624-1845) | 276 (242-297) |
| 5.4410 | 25,570 (0.51) | 8.11% | 99.50% | 1.11% | 90.0 | 16.1 | 1317 (1174-1440) | 278 (222-304) |
| 4.6181 | 5,257 (0.10) | 3.14% | 99.90% | 2.09% | 47.8 | 30.2 | 1371 (1180-1562) | 304 (218-366) |
| AUC at 28m 0.740 (0.732,0.748), risk=0.07015% | | | | | | | | |
| 7.8045 | 759,791 (15.02) | 45.18% | 85.00% | 0.21% | 474.0 | 3.0 | 2162 (2035-2274) | 300 (285-319) |
| 7.2962 | 557,358 (11.02) | 38.30% | 89.00% | 0.24% | 410.1 | 3.5 | 2070 (1955-2181) | 292 (274-308) |
| 6.6809 | 253,616 (5.01) | 24.75% | 95.00% | 0.35% | 288.9 | 4.9 | 1894 (1805-2043) | 284 (263-305) |
| 6.4077 | 152,348 (3.01) | 18.57% | 97.00% | 0.43% | 231.2 | 6.2 | 1748 (1639-1854) | 279 (247-303) |
| 5.4410 | 25,570 (0.51) | 8.09% | 99.50% | 1.12% | 89.1 | 16.0 | 1347 (1190-1447) | 278 (226-306) |
| 4.6181 | 5,257 (0.10) | 3.18% | 99.90% | 2.15% | 46.5 | 30.6 | 1396 (1202-1572) | 309 (221-374) |
| AUC at 29m 0.741 (0.733,0.749), risk=0.07111% | | | | | | | | |
| 7.8045 | 759,806 (15.02) | 45.43% | 85.00% | 0.22% | 465.0 | 3.0 | 2173 (2043-2282) | 306 (289-325) |
| 7.2963 | 557,376 (11.02) | 38.45% | 89.00% | 0.25% | 403.0 | 3.5 | 2074 (1974-2185) | 297 (279-318) |
| 6.6809 | 253,618 (5.01) | 24.85% | 95.00% | 0.35% | 283.7 | 5.0 | 1914 (1818-2063) | 289 (267-310) |
| 6.4077 | 152,348 (3.01) | 18.57% | 97.00% | 0.44% | 228.1 | 6.2 | 1766 (1658-1859) | 284 (257-306) |
| 5.4410 | 25,570 (0.51) | 8.15% | 99.50% | 1.15% | 87.3 | 16.1 | 1351 (1209-1454) | 283 (236-308) |
| 4.6181 | 5,257 (0.10) | 3.20% | 99.90% | 2.19% | 45.7 | 30.8 | 1371 (1218-1562) | 319 (231-379) |
| AUC at 30m 0.741 (0.733,0.749), risk=0.07135% | | | | | | | | |
| 7.8045 | 759,819 (15.02) | 45.44% | 85.00% | 0.22% | 463.3 | 3.0 | 2173 (2043-2282) | 307 (291-325) |
| 7.2963 | 557,380 (11.02) | 38.49% | 89.00% | 0.25% | 401.3 | 3.5 | 2074 (1974-2185) | 299 (281-319) |
| 6.6809 | 253,621 (5.01) | 24.88% | 95.00% | 0.35% | 282.4 | 5.0 | 1914 (1818-2063) | 292 (270-311) |
| 6.4077 | 152,348 (3.01) | 18.62% | 97.00% | 0.44% | 226.7 | 6.2 | 1766 (1658-1859) | 286 (258-307) |
| 5.4410 | 25,570 (0.51) | 8.15% | 99.50% | 1.15% | 87.0 | 16.1 | 1351 (1209-1454) | 284 (236-310) |

| | | | | | | | | |
|---|---|---|---|---|---|---|---|---|
| 4.6181 | 5,257 (0.10) | 3.21% | 99.90% | 2.21% | 45.3 | 30.9 | 1371 (1218-1562) | 320 (231-385) |
